# Physical activity interventions for women with metastatic breast cancer: A systematic review of published and ongoing randomised controlled trials

**DOI:** 10.1101/2025.08.06.25333137

**Authors:** Louise H. Hall, Sophie M.C. Green, Zainab Haider, Abigail Fisher, Natalie King, Rebecca J Beeken, Samuel G. Smith

**Author notes:** Contributed equally to the work and should be considered joint senior authors (Rebecca J Beeken, Samuel G. Smith). Corresponding author Professor Samuel G. Smith University of Leeds, Leeds Institute of Health Sciences Clarendon Way, LS2 9NL.

## Abstract

**Purpose:** We systematically reviewed published and ongoing physical activity (PA) trials in women with metastatic breast cancer (MBC). We examined: i) the effectiveness of PA interventions on key outcomes, and identified: ii) the type of interventions being evaluated, iii) how they were delivered; iv) their theoretical basis.

**Methods:** Seven databases and two trial registries were searched in August 2024 for; randomised controlled trials, testing any PA intervention in people with MBC, reporting a PA outcome. The Joanna Briggs Institute (JBI) handbook was followed, including quality assessment using the JBI Critical Appraisal Checklist for RCTs. Data were summarised narratively. Intervention details were extracted using the TIDieR framework.

**Results:** 1687 records were screened and 96 assessed for eligibility. 28 reports were included (13 full reports, 4 protocols, 11 trial registries). Sample sizes ranged from 21-357 participants. 21/28 reports were phase II, pilot, or feasibility trials. Most interventions did not cover all types of recommended PA. Methodological quality of studies was moderate. Intervention adherence was moderate to high (≥50% in 10 studies). Among studies reporting on safety (9), only one recorded any serious events (two events) related to the intervention. Evidence indicates that PA can improve fatigue, health-related QoL, physical fitness and functioning over the short and medium-term (≤6 months).

**Conclusions:** Physical activity is safe, well adhered to, and improves physical function and QoL in MBC. Future trials could clarify the optimal PA type, duration, delivery mode, frequency, and long-term effectiveness. Women with MBC should be supported by healthcare professionals to be active.

## Introduction

Treatment advances in metastatic breast cancer (MBC) have improved survival across all disease sub-types^[1]^. There were an estimated 167,518 prevalent cases of metastatic breast cancer (MBC) in the US in 2020, a figure projected to rise to 246,194 by 2030^[2]^. A quarter of this growing population will live over five years following diagnosis^[3]^. The medical and productivity costs of MBC is estimated to be $63.4B in 2015, increasing 140% to $152.4B by 2030^[2]^. These costs are expected to be higher for younger and midlife women^[4]^.

Maintaining functional quality of life (fQoL) is a priority for patients, but women currently undergo multiple lines of treatment, causing a wide range of short- and long-term physical and psychological consequences including reduced physical function, fatigue, pain, and distress^[5–10]^. Women experiencing severe symptoms are more likely to cease treatment early, which may impact survival^[11]^. While medical intervention may be required for some of these physical and psychological sequelae of treatment, effective self-management of mild to moderate symptoms could improve patient wellbeing and clinical outcomes.

Definitive evidence shows physical activity (PA) can improve fQoL in early stage breast cancer^[12]^, and it is recommended in major guidelines (e.g. American Society of Clinical Oncology)^[13,14]^. Although women with MBC are often excluded from PA trials due to safety concerns and historically poor prognosis, PA is considered safe^[15,16]^, including for those with bone metastases when modified by a trained professional^[17–19]^. Furthermore, observational evidence suggests PA could improve fQoL in women with MBC^[20]^. Improving fQoL should be prioritised, as across a range of disease sites and stages higher fQoL is associated with lower emergency admissions and hospitalisations ^[21]^, reduced sick leave^[22,23]^, and in MBC potentially longer survival^[24–26]^.

Existing systematic reviews of PA interventions have included trials enrolling people with metastatic cancer across all disease sites, but few trials enrolled MBC patients^[27–29]^. While some PA interventions may be applicable to all disease sites, specific modifications may be required for MBC^[30]^, and generalisations from other cancer sites should not be made without evidence of safety, efficacy, and feasibility in this patient group. A systematic synthesis of physical activity trials specifically in MBC is needed to inform the development of future intervention strategies. It could also provide a single resource of trials within this field for healthcare professionals (HCPs) wanting to implement PA in their clinical practice. We aimed to systematically synthesise the literature on published and ongoing PA trials in women with MBC. Our objectives were to examine: i) the effectiveness of physical activity interventions, and to identify: ii) the types of physical activity interventions being evaluated, iii) how they are delivered; and iv) the theoretical basis for them.

## Method

The review was conducted following the Joanna Briggs Institute’s Manual for Evidence Synthesis^[31]^ and reported using the Preferred Reporting Items for Systematic Reviews and Meta-Analyses (PRISMA:2020)^[32]^ (Appendix 1). It was pre-registered on PROSPERO (CRD42023462994).

### Search strategy

Searches for completed randomised controlled trials (RCTs) and published protocols of ongoing trials were conducted on 16^th^ August 2024 in MEDLINE, Cochrane, CINAHL, PSycINFO, Web of Science, Scopus and Embase. Ongoing trials were searched for on clinicaltrials.gov and ICTRP on 20th August 2024. There were no restrictions on date. Keywords were identified from known papers on similar topics. An information specialist (NK) created our search strategy for MEDLINE and reviewed our search strategies for all other databases. See Appendix 2 for search terms for MEDLINE, and the Open Science Framework (OSF) [DOI 10.17605/OSF.IO/2SW76] for search terms for all databases. Keywords and MESH terms were searched for: ((metastatic OR advanced) breast cancer) AND (physical activity OR exercise OR weight training OR resistance training), with filters for randomised controlled trials applied.

### Eligibility criteria

Studies were included if they used an RCT design (including pilot and feasibility trials), were available in English, and met the following PICO criteria: Population: adults (aged 18+) with any type of advanced or MBC; Intervention and Context: any type of physical activity intervention delivered in any suitable setting; Comparator: usual care or a comparable intervention or any other suitable comparator; Outcome: measure of physical activity. Additional outcomes of interest included Quality of Life, Fatigue, Intervention Adherence, Adverse Events. Full eligibility criteria can be found on the OSF.

### Study screening and selection

Completed trials and published protocols were downloaded into Endnote and duplicates removed, before uploading onto Rayyan for screening. Titles and abstracts were independently screened in duplicate by RB, SS, LH. Full-texts were independently screened in duplicate by SS and LH. Discrepancies were arbitrated by RB and SG. Forward citation searching and hand searching reference lists of included studies was conducted. Ongoing trials were downloaded into Microsoft Excel and duplicates removed before titles, abstracts, and full texts/registry entries were independently screened in duplicate by RB and LH, with discrepancies arbitrated by SS.

### Assessment of methodological quality

Results papers were independently assessed for methodological quality using the Joanna Briggs Institute Critical Appraisal Checklist for RCTs^[33]^ in duplicate by RB, AF and SS, arbitrated by SG. As the majority of studies were pilot and feasibility trials, they were appraised at the study level rather than the outcome level. Phase III trials were assessed for the primary outcome only.

### Data extraction

Data was extracted in duplicate by LH and SS, (arbitrated by SG and ZH), in Microsoft Excel. The form collected data on; study design, country, sample size, key clinical eligibility criteria, locations of metastases, disease sub-types, relevant treatment information, participant demographics, socio-economic variables, outcome measures and timepoints, patient-reported outcomes assessed, biomarkers assessed, effect sizes of primary and secondary outcomes of interest, intervention adherence, recruitment rate, retention, and adverse events. Intervention details were extracted using the template for intervention description and replication (TIDieR) framework^[34]^.

### Data synthesis

We undertook a narrative synthesis structured around type of report (results, protocol, registry), study characteristics, types of outcome measures, intervention characteristics, and study findings. Summaries of the interventions are tabulated using the extracted data, and key information described in text. Commonalities and differences between the overall populations of studies are identified and described. No meta-analysis was planned, as initial searches indicated high heterogeneity of content, context, and outcome measures.

LH and SS categorised the interventions as containing or not containing the following types of exercise: 1) Aerobic; 2) Resistance / Strength Training; 3) Flexibility, Stretching, Mobility; 4) Balance, Functional; 5) Other. These categories were based on WHO exercise recommendations^[35]^, Harvard Medical School descriptions of important exercise types^[36]^, a Taxonomy of PA interventions in Older Adults^[37]^, and a systematic review that categorised types of PA interventions^[38]^.

## Results

The flow of full reports, protocol papers and trial registry entries are shown in Figure 1. The database searches yielded 1795 records. After removing duplicates, 1687 records were screened and 96 were assessed for eligibility. Seventy-one records were excluded, leaving 25 reports for inclusion. After reviewing records known to the authors and completing backwards and forwards citation searching, an additional 3 reports were included. In total, 28 reports were included, including 13 full reports^[39–51]^, 4 protocol papers^[52–55]^ and 11 trial registry entries^[56–66]^ (Table 1).

**Figure 1.**
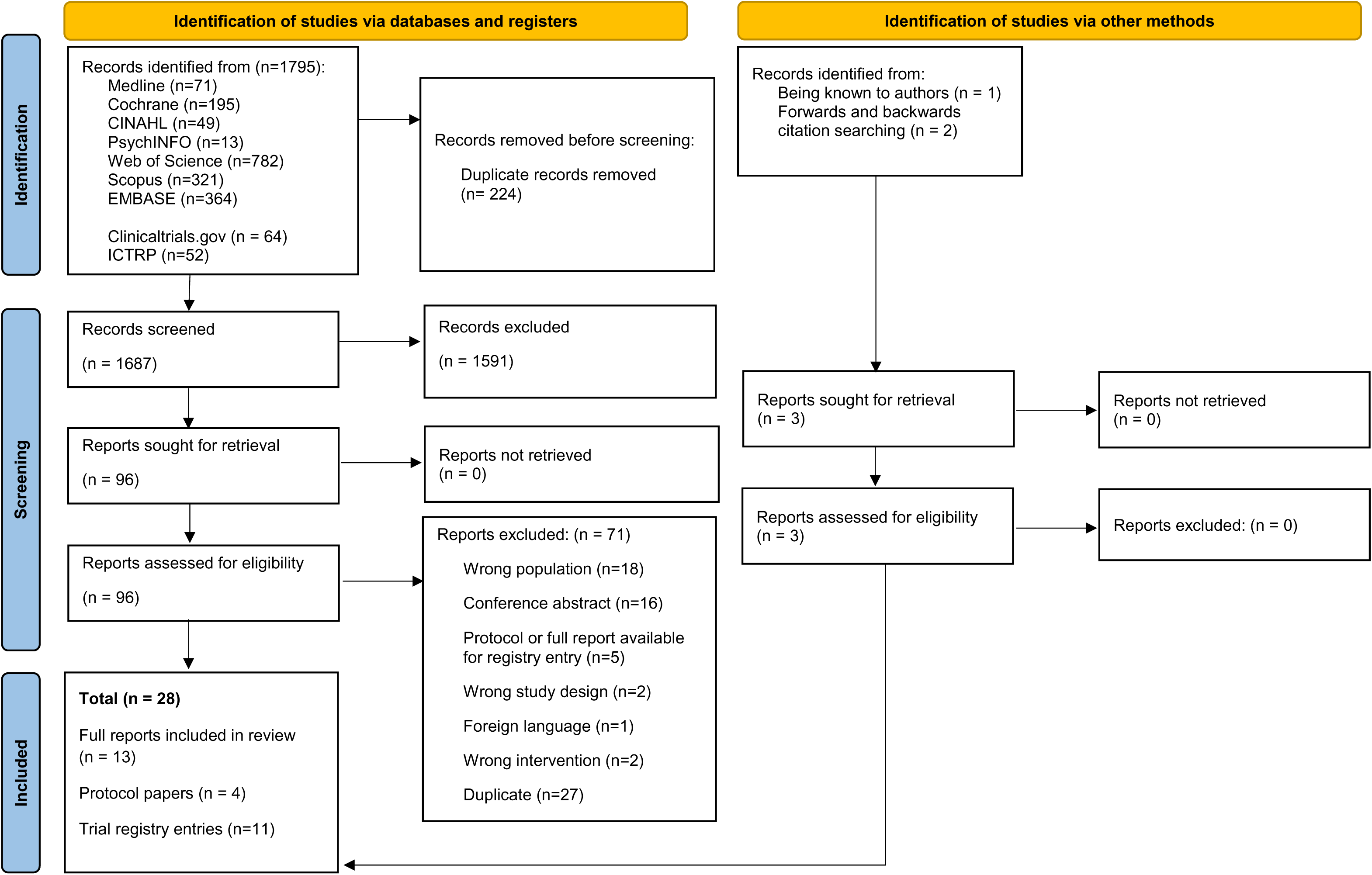
PRISMA flow chart.

**Table 1.** Summary of characteristics in full reports, protocols and trial registries.

| Authors / Date | Country | Design | Sample size | Follow-up | Key eligibility criteria | Intervention / comparator | Primary outcome(s) | Participant characteristics | Findings | Safety |
| --- | --- | --- | --- | --- | --- | --- | --- | --- | --- | --- |
| Full reports |  |  |  |  |  |  |  |  |  |  |
| Carson, 2021<sup>[39]</sup> | USA | Pilot, RCT (2:1) | 63 | Daily diaries, 8 weeks | MBC. Excluded ECOG $\geq 3$ or KPS $\leq 50$, current engagement in yoga practice 1 or more days per week. | Yoga vs. support group | Daily pain (0-9 self-report) | Mean age: 57.5; 74.5% White; 58.3% bone metastases; 14.6% brain metastases | No intervention x time interaction, suggesting no effect on daily pain (p=0.275) | Not reported |
| Headley, 2004<sup>[40]</sup> | USA | Pilot, quasi-RCT | 38 | 12 weeks | MBC, scheduled to initiate outpatient chemotherapy, performance status $\leq 2$, ability to sit on straight backed chair for 30 minutes. Excluded if receiving high dose chemotherapy or receiving only hormonal therapy | Seated exercise vs. usual care | Fatigue (FACIT-F) | 51 years, 84.4% White | Intervention group had less decline in scores on fatigue subscale (p=.0078) | No safety events related to intervention |
| Hiensch, 2024<sup>[50]</sup> | Germany, Netherlands, Poland, Spain, Sweden, Australia | Phase III, RCT | 357 | 3, 6, 9 months | MBC, ECOG $\leq 2$. Excluded unstable bone metastases, untreated brain metastases, and current engagement in intense exercise training | Multimodal: aerobic, resistance and balance + physical activity trackers vs. usual care + physical activity tracker + exercise advice | Fatigue (EORTC QLQ FA12) and health related QoL (EORTC QLQ C30) | 99.4% female; 55.4 years; Treatment: endocrine therapy (52.3%); targeted therapy (57.1%); chemotherapy (25.5%); bone-modifying agent (46.5%). Metastases locations: Bone (67.2%); lung (26.6); liver | Recruitment rate = 42%, Attrition at 6 months = 18.5% Exercise group had sig. improvements in HRQoL at 6 months (ES = 0.33, p=.0003, BGD = 4.8 (95% CI 2.2-7.4), lower | 2 exercise related SAEs unrelated to bone mets (wrist fracture; sacral stress fracture). 80 AEs including pain (51.3%), dizziness (12.5%), muscle soreness and/or cramps (11.3%), fatigue (8.9%); requiring modifications to |
| | | | | | | | | (34.7%); lymph node (38.7%). Triple negative (9.8%), HER2positive (23.2%), HER2negative/HR positive (59.7%) | physical fatigue (ES = 0.22, $p = 0.27$, BGD = -5.3, 95% CI = -10 to -0.6). Sig. improvements seen for physical fitness and various QLQ-C30 subscales. | the exercise programme |
| Ligibel, 2016<sup>[41]</sup> | USA | Phase not described, RCT (1:1) | 101 | 16 weeks | MBC or locally advanced disease; ECOG performance status 0 or 1; baseline performance of $\leq 150$ mins weekly recreational physical activity. Excluded women with untreated brain metastases. | Aerobic exercise vs. wait-list control | Physical function: EORTC QLQ C30 and Bruce Ramp Treadmill Test | Median age: 49 years; 94% White; Treatment: 53% endocrine therapy, 38.7% chemotherapy, 36.7% biologic therapy, 3% no treatment; 66% visceral metastatic disease | Non-significant improvement in physical function using EORTC QLQ C30 ($p=0.23$) Non-significant increase in exercise duration using Bruce Ramp Treadmill Test ($p=0.35$). Intervention participants increased their minutes of exercise by a mean of 62.4 ± 102.8 minutes per week versus | No safety events occurred |
|  |  |  |  |  |  |  |  |  | 46.0 +/- 154.3 minutes per week among controls (P = .17). |  |
| Oh, 2014<sup>[42]</sup> | Australia | Phase II (feasibility), RCT | 27 | 5 and 10 weeks | MBC; ECOG 0-2. Excluded women with brain metastases and already practicing Qigong / meditation once a week in last 6 months | Qigong vs. meditation | Adherence to programme (in person, home practice) and adverse events | 57.4 years; 78.6% (intervention)<br>92.3% (control)<br>White | 63% completed study, 80% adhered to intervention, unknown proportion completed home practice 3-4 days/week for 15-30 minutes per day. | No safety events occurred |
| Phillips, 2024<sup>[51]</sup> | USA | Pilot / Feasibility RCT | 49 | 12 weeks | MBC, low levels of physical activity. Excluded untreated brain metastases. | App-based PA promotion to promote walking and behaviour change + FitBit + weekly coaching calls vs. healthy lifestyle attention control | Feasibility: ≥ 80% completion rate (# complete 12-week assessment / # randomised); Adherence: % days wearing FitBit; Acceptability: Satisfaction | 54.8 years; 85.7% white, 14.3% non-white (6.1% Black, 2.0% Asian pacific islander, 6.1% other or unknown). Hispanic (n=5, 10.2%). Current treatment: 55.1% chemotherapy; 10.2% radiation therapy; 47.9% immunotherapy; 12.5% targeted therapy; 53.1% hormone therapy; | Completion rate = 98%<br>Intervention group wore the Fitbit 92.7% of the time, met their step goal 53.4%, set a step goal 87.4%, used the app 94.1% of study days. ITT showed significant effect favouring intervention group on self-efficacy ($p =$ | 1 death. No other Adverse Events were reported. |
| | | | | | | | | | .03), PA goal setting ($p < .01$). Trends ($d = .025$ to $0.45$) for improvements on physical activity, (daily steps, activity counts, MVPA minutes, smaller declines in total PA and light PA, smaller increases in sedentary time). | |
| Porter, 2019<sup>[43]</sup> | USA | Pilot, RCT (2:1) | 65 | 3 and 6 months | MBC. Excluded women if ECOG 3+, KPS <60; currently practicing yoga 1+ day/week | Yoga vs. social support | Acceptability (Client Satisfaction Questionnaire); Feasibility (session attendance and assessment completion) | 57.3 years; White (74.6%); Treatment: radiotherapy (67.7%), surgery (24.6%), chemotherapy ongoing (64.6%) | 82% indicated high satisfaction with intervention; 65% attended $\geq 4$ sessions Intervention group walked a greater distance at post-intervention and 6-months (6-min walk test). | No safety events related to intervention |
| Schmitz, 2023<sup>[44]</sup> | USA | Pilot / feasibility, RCT with partial crossover | 42 | 3 months | MBC. Excluded women if ECOG >2. | mHealth vs. waiting list control | Acceptability (% agreeing to participate); Feasibility (% interacting with platform at least 30/90 days) | 53.4 years; White (90.5%) | 51% consent rate for those known to be eligible; 64.9% logged in 30 of the 90 days; adherence declined each month after weekly calls; No BGD in sit-stand test. | No safety events occurred |
| Schmitz, 2021<sup>[45]</sup> | USA | Pilot / feasibility, RCT with partial crossover | 21 | 3-months | MBC, ECOG $\leq 3$ | mHealth vs. waiting list control | Feasibility (Proportion consenting, interacting with platform and participating in at least one month of programme) Acceptability (Proportion of eligible patients agreeing to participate) | 60 years; 94% White Treatment: No anti-cancer treatment (11.8%); systemic infusion therapy (70.6%); hormone therapy (64.7%); CDK 4/6 inhibitors (41.2%). | Feasibility: 48.3% (40.3/90 logged into Nurse AMIE. 13/17 (76%) who actively participated were still using at 1 month). Acceptability: 68% Signal of efficacy: No group x time interaction for pain, fatigue, distress or sleep. | No additional healthcare visits incurred as a result of the intervention |
| Scott, 2018<sup>[46]</sup> | USA | Pilot / Feasibility RCT | 65 | 3-months | MBC, ECOG 0 or 1, performance of <150mins of moderate-intensity exercise per week | Supervised aerobic training vs. supervised stretching | Feasibility (loss to follow-up and attendance) | 54 years. Treatment: Current therapy (chemotherapy (57%); endocrine therapy (48%); | Loss to follow-up (3%); Mean attendance (63% ± 30%); Permanent discontinuation | Aerobic training group: 36% stopped a session early due to nonserious health event. 73% |
| | | | | | | | | radiotherapy (5%); prior lines of therapy (0=12%; 1=40%; 2=31%; $\geq 3$ = 35%). | of aerobic training (27%); 12% of sessions required a dose reduction. No BGD on functional capacity (6-min walk test, sit-stand test). | experienced at least 1 nonserious adverse event. No serious adverse events observed. Attention control group: 7 nonserious adverse events |
| Sheean, 2021<sup>[47]</sup> | USA | Pilot / Feasibility Crossover RCT | 40 | 12-weeks | MBC; ECOG 0-1, no change in anti-neoplastic therapies (30 days), no severe pain (≥ grade 3 NCI CTCAE), currently non-compliant with ACS nutrition and physical activity guidelines | Multimodal diet and physical activity vs. waitlist control | Feasibility (100% of recruitment goal)<br>Adherence ($>75\%$ telephone sessions and 2/3 exercise sessions)<br>Efficacy (trends in improvement of QoL)<br>Safety events | 55 years, non-Hispanic White (91.4%); Current treatment: endocrine (45.2%); chemotherapy (25.8%); immunotherapies (71.0%) | Attendance: 86% (93% in-person and 84% telephone sessions)<br>Significant improvements in Fact G ($p=0.003$) with improvements in physical, emotional and functional wellbeing;<br>Increases in handgrip strength ($p=0.003$) and time spent in physical activity ($p<0.001$) | No adverse events related to the intervention; three women (2 Intervention, 1 Waitlist control) developed new symptoms/disease progression |
| Vadiraja, 2017<sup>[48]</sup> | India | Phase not described, RCT | 91 | 3-months | MBC; 30-70 years; ambulatory $>50\%$ of time. | Yoga vs. supportive therapy and education | Not described | 50.5 years; | Improvements in fatigue severity, frequency, | Not reported |
|  |  |  |  |  | Excluded women with brain mets and who underwent chemotherapy |  |  |  | interference and diurnal variation (p<0.0001) |  |
| Yee, 2019<sup>[49]</sup> | Australia | Pilot / Feasibility RCT | 14 | 8-, 16 weeks | MBC; ECOG 0-2. Excluded women who regularly participated in physical activity | Supervised resistance training and unsupervised walking vs. usual care | Feasibility (recruitment, retention, adherence, safety) | 62.2 years; | Recruitment rate = 93%; retention rate = 100% [intervention] / 83% [usual care]. 100% attendance to 16 supervised exercised sessions; all completed at prescribed intensity and volume. 2/8 (25%) women adhered to the walking program; compliance to walking intensity=71%. Improvements shown in fatigue (FACIT-F ES =1.92), pain (ES= 1.61), quality of life (physical, role, emotional, social function), and physical assessments. | No safety events occurred |
| Protocols |  |  |  |  |  |  |  |  |  |  |
| Delrieu, 2020<sup>[52]</sup> | France | Phase III, RCT | 244 | HRQoL= 6,12,18,36,48 weeks, month 18<br>Other = months 3,6,12,18 | MBC with at least one positive hormone receptor (HR+) and HER2- | Walking (app and tracker) vs. physical activity recommendation | Time to deterioration of global health status and fatigue (EORTC QLQ C30) | N/A | N/A | N/A |
| Hart, 2018<sup>[53]</sup> | Australia | Pilot / Feasibility RCT | 40 | 13 weeks | Stable osteolytic bone metastases in verval, thoracic, and/or lumbar vertebrae; have not engaged in regular exercise in previous 3 months | Multimodal (resistance, aerobic and flexibility) focusing on spinal isometric training vs. usual care | Feasibility: recruitment, completion, safety, programme adherence and compliance. FACIT. | N/A | N/A | N/A |
| Kirkham, 2021<sup>[54]</sup> | Canada | Phase II, RCT | 50 | pre cycle 1, pre cycle 3 post cycle 6 and 2-years | MBC, starting IV chemotherapy, ECOG $\leq 2$ | Cycling and dietary change during chemotherapy Vs. Usual care PLUS one-time phone consultation with a registered dietitian and exercise physiologist vs. usual care plus one nutrition and exercise advice phone call | Change in lesion size (CT scan) | N/A | N/A | N/A |
| Meier-Girard, 2020<sup>[55]</sup> | Switzerland | Phase III, RCT | 196 | 8, 14 and 20 weeks | MBC, FACIT-F score <34, ECOG ≤1 | Eurythmy + CoordiFit vs. CoordiFit only | FACIT-F subscale | N/A | N/A | N/A |
| Trial registries |  |  |  |  |  |  |  |  |  |  |
| Ballinger, 2022<sup>[56]</sup> | USA | Phase II, RCT | 100 | 8- and 16 weeks | MBC, ECOG 0-2, Excluded: women receiving cytotoxic chemotherapy in past 12 months, active untreated brain metastases | Multimodal (cardiovascular, resistance + balance / stretching) vs. usual care plus leaflet | Cardiorespiratory fitness (minutes on treadmill) | N/A | N/A | N/A |
| Borquist, 2024<sup>[66]</sup> | Denmark | Feasibility RCT | 21 | 1 year, 1.5 years | MBC, BMI ≥ 25, Receive first-line endocrine-based therapy, ECOG 0-1 | Aerobic, resistance, stretching vs. | Recruitment rate | N/A | N/A | N/A |
| Chamberlin, 2012<sup>[57]</sup> | USA | Pilot / Feasibility RCT | 4 | 2, 3, 4, 5, 6- months | MBC, Karnofsky performance status ≥80%; excluded women with untreated CNS disease | Walking vs. usual care | Physical activity / steps (exercise logs and pedometer) | N/A | N/A | N/A |
| Crane, 2024<sup>[65]</sup> | USA | Phase III, RCT | 260 | 3, 6, 12 months | ER+ and/or PR+, HER-2 metastatic or locally advanced BC; excluded unstable bone metastases, unstable cardiac disease, oxygen dependant, not consistently meeting exercise guidelines in past 3 months. | Exercise (aerobic + resistance) vs. exercise + fasting vs. fasting only vs attention control (bi-weekly remote health education) | Fatigue (EORTC QLQ C30) | N/A | N/A | N/A |
| Dal Molin, 2022<sup>[64]</sup> | Brazil | Phase not described, RCT | 68 | 2, 4, 6 months |  | App + SmartWatch nutrition and physical activity vs. App only | Quality of Life (FACT-B) | N/A | N/A | N/A |
| Devoto, 2014<sup>[58]</sup> | Australia | Phase not described, RCT | 48 | Weeks 1, 4, 6, and 14-21 days post week 6 | MBC, experiencing fatigue, ECOG 0-2; excluded: recent or anticipated change to systemic therapy, bone metastases with lytic metastatic lesions involving long bones | Multimodal (Aerobic and resistance) vs. active control (undefined) | Fatigue (FACIT-F) | N/A | N/A | N/A |
| Harvie, 2015<sup>[59]</sup> | UK | Phase II, RCT | 134 (planned) 68 (final) | Day 1 of each chemotherapy cycle, plus every 9-12 weeks during treatment. 15 month follow up |  | Resistance training + Diet vs. resistance training alone | Progression free survival | N/A | N/A | N/A |
| Liu, 2020<sup>[60]</sup> | Australia | Pilot / Feasibility, RCT | 20 | 12 and 18 weeks |  | Behavioural Counselling vs. attention control (physical activity recommendation, handout, FitBit and diary) | Physical Activity (Actigraph Accelerometer and IPAQ=SF) | N/A | N/A | N/A |
| Lozano-Lozano, 2022<sup>[61]</sup> | Spain | Phase not described, RCT | 58 | 14 weeks |  | Aerobic vs. usual care | Functional capacity (6 minute walk test) and Quality of life (EORTC QLQ C30) | N/A | N/A | N/A |
| Reeves, 2022<sup>[62]</sup> | Australia | Phase III, RCT | 200 | 3, 6, 9, 12 months |  | Aerobic, resistance exercises + diet vs. usual care + encouragement to access services | Physical function (EORTC QLQ C30) and whole body lean mass (DXA body scan) | N/A | N/A | N/A |
| Somayeh Nazari, 2023<sup>[63]</sup> | Iran | Phase not described, RCT | 28 | 8 weeks |  | Resistance training vs. no intervention | Liver enzymes ALT, AST and ALP. | N/A | N/A | N/A |

RCT= Randomised controlled trial. MBC = Metastatic Breast Cancer. ECOG = Eastern Cooperative Oncology Group. KPS= Karnofsky Performance Scale. FACIT-F= Functional Assessment of Chronic Illness Therapy-Fatigue. EORTC QLQ C30= European Organization for the Research and Treatment of Cancer Quality of Life Questionnaire. CDK= cyclin-dependent kinase. NCI CTCAE= National Cancer Institute Common Terminology Criteria for Adverse Events. ACS= American Cancer Society. QoL= Quality of life. HRQoL= Health Related Quality of Life. IPAQ-SF= International Physical Activity Questionnaire- Short Form. DXA= Dual-Energy X-ray Absorptiometry. ALT= alanine transaminase. AST= aspartate transaminase. ALP= alkaline phosphate. ES = Effect Size. BGD = Between Group Difference.

### Full reports

#### Quality assessment

Table 2 summarises the methodological quality of full reports. True randomisation was used in all except two studies, from the same authors, which lacked detail about the randomization procedure^[44,45]^. Those allocating participants to groups were concealed to allocation in eight studies^[39,43,46–51]^, but it was unclear in the remaining five^[40–42,44,45]^. As expected for behavioural interventions, no study blinded participants or intervention deliverers to treatment allocation. Outcome assessors were not blinded to treatment allocation in two studies^[49,50]^, and this was unclear in another six^[40,42,44–46,48]^.

**Table 2.** Summary of quality assessments using the Joanna Briggs Institute Critical Appraisal Checklist for Randomized Controlled Trials.

|  | True randomisation? | Allocation concealed? | Groups similar? | Participants blinded? | Intervention delivery blinded? | Outcome assessors blinded? | Groups treated identically? | Follow-up complete? Differences | Analysed within randomised group? | Outcomes measured identically? | Outcomes reliable? | Appropriate statistics? | Design appropriate? |
| --- | --- | --- | --- | --- | --- | --- | --- | --- | --- | --- | --- | --- | --- |
| Carson<sup>[39]</sup> | yes | yes | yes | no | no | yes | no | yes | yes | yes | yes | yes | yes |
| Headley<sup>[40]</sup> | yes | unclear | no | no | no | unclear | yes | yes | yes | yes | yes | yes | yes |
| Hiensch<sup>[50]</sup> | yes | yes | yes | no | no | no | yes | yes | yes | yes | yes | yes | yes |
| Ligibel<sup>[41]</sup> | yes | unclear | yes | no | no | yes | yes | yes | yes | yes | yes | yes | yes |
| Oh<sup>[42]</sup> | yes | unclear | yes | no | no | unclear | no | no | yes | yes | yes | yes | yes |
| Phillips<sup>[51]</sup> | yes | yes | unclear | no | no | yes | unclear | yes | yes | yes | yes | yes | yes |
| Porter<sup>[43]</sup> | yes | yes | no | no | no | yes | no | no | yes | yes | yes | yes | yes |
| Schmitz<sup>[44]</sup> | unclear | unclear | no | no | no | unclear | no | no | yes | yes | yes | no | yes |
| Schmitz<sup>[45]</sup> | unclear | unclear | yes | no | no | unclear | unclear | no | yes | yes | yes | yes | yes |
| Scott<sup>[46]</sup> | yes | yes | yes | no | no | unclear | yes | yes | yes | yes | yes | yes | yes |
| Sheean<sup>[47]</sup> | yes | yes | yes | no | no | yes | no | no | yes | yes | yes | yes | yes |
| Vadiraja<sup>[48]</sup> | yes | yes | unclear | no | no | unclear | no | no | yes | yes | yes | yes | yes |
| Yee<sup>[49]</sup> | yes | yes | yes | no | no | no | yes | yes | yes | yes | yes | yes | yes |

Baseline differences existed between groups in three studies^[40,43,44]^; including differences in education level^[40,43]^, marital^[40]^ and employment statuses^[44]^. The intervention and control groups were treated differently in six studies^[39,42–44,47,48]^. For example, offering the control group support unavailable to the intervention group, such as enhanced usual care^[42]^ or additional communication^[47]^.

Differences between groups regarding follow-up (e.g., descriptions of; loss to follow-up (LTF), reasons for LTF, impact of incomplete follow-up) were inadequately described in six studies^[42–45,47,48]^. All studies had reliable outcomes that were measured identically across groups, used appropriate designs and analysed participants in the groups they were randomized. One study was considered to not have used appropriate statistical analysis^[44]^, which involved a complex crossover design but conducted separate comparisons for the immediate and delayed receipt of the intervention with the control.

#### Characteristics of studies

Most (61.5%) full reports originated in the USA^[39–41,43–47,51]^, with representation also from Australia^[42,49]^, India^[48]^, and various European countries^[50]^. Three were full reports of phase III trials^[41,48,50]^, with the remainder being phase II^[42]^ or pilot/feasibility studies (69.2%)^[39,40,43–47,49,51]^. Nearly all used a parallel group RCT design, with a minority using an RCT with full or partial cross-over^[44,45,47]^. One trial reportedly used a quasi-experimental design, but it was unclear how the allocation method was not random^[40]^. Sample sizes ranged from 21^[45]^ to 357^[50]^ (median=49).

Participant characteristics were not consistently reported, with 9/13 reporting on ethnicity^[39–45,47,51]^, and 9/13 reporting any socio-economic variable^[39,40,42–44,47,49–51]^. Of these, participant samples were primarily White (74.5-94.0%) with the average age ranging from 49^[41]^ to 62.2^[49]^ years. Of the seven studies reporting employment status, the percentage of participants working full- or part-time ranged from 28.6%^[43]^ to 61.2%^[51]^. Eight papers reported on recruitment rates, which ranged from 21.1%^[51]^ to 93.0%^[49]^ (mean=49.8%).

Disease sub-type, treatment information, and location of metastases were not always reported. Where they were, bone metastases (29-67.2% of participants, reported in 4 studies^[39,47,49,50]^) and visceral metastases (up to 71% reported in 6 studies^[39,41,46,47,49,50]^) were common. Seven studies included participants with an Eastern Cooperative Oncology Group (ECOG) rating of ≤2^[39,40,42–44,49,50]^, one included those with ECOG of ≤3^[45]^, and three with an ECOG of ≤1^[41,46,47]^.

Primary outcome measures were; self-reported pain^[39]^, fatigue^[40]^, quality of life^[50]^, feasibility and acceptability^[42–47,49,51]^. One trial had co-primary outcomes of physical function assessed by the EORTC QLQ C30 and the Bruce Ramp Treadmill Test^[41]^. One trial did not specify their primary outcome^[48]^. Two reports listed safety as a primary outcome^[47,49]^.

Follow-up ranged from 8 weeks^[39]^ to 9 months^[50]^. Four full reports used a wait-list comparator^[41,44,45,47]^, two used usual care^[40,49]^, and seven^[39,42,43,46,48,50,51]^ used a variety of approaches to enhanced usual care (e.g. attention controls^[46,51]^, support groups^[39,43]^).

#### Intervention characteristics according to the TIDieR checklist

Table 3 summarises the intervention characteristics according to the TIDieR checklist. Appendix 3 summarises the full data extraction of the TIDieR components. Three full reports referred to a theory or theoretical framework when describing the intervention^[40,47,51]^: two were based on social cognitive theory^[47,51]^, but one of these was not justified or described in detail^[47]^. One study reported using Roy’s Adaptation Model as a conceptual framework to guide their intervention of a seated exercise program^[40]^. One further study, investigating a mindful yoga-based intervention, described their rationale as increasing psychological processes (e.g., acceptance, mindfulness) to improve cancer related pain, fatigue, and distress^[39]^.

**Table 3.**
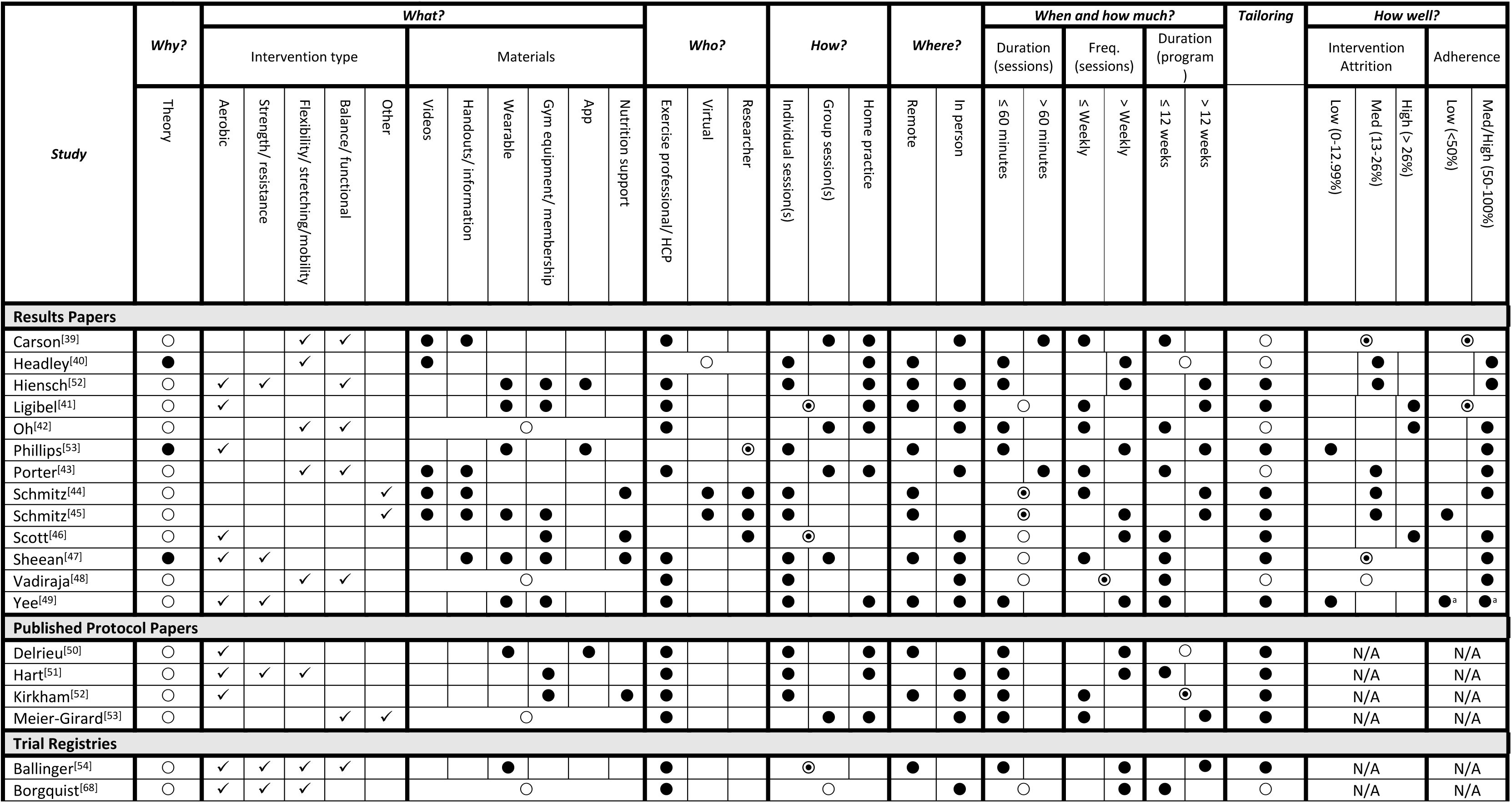
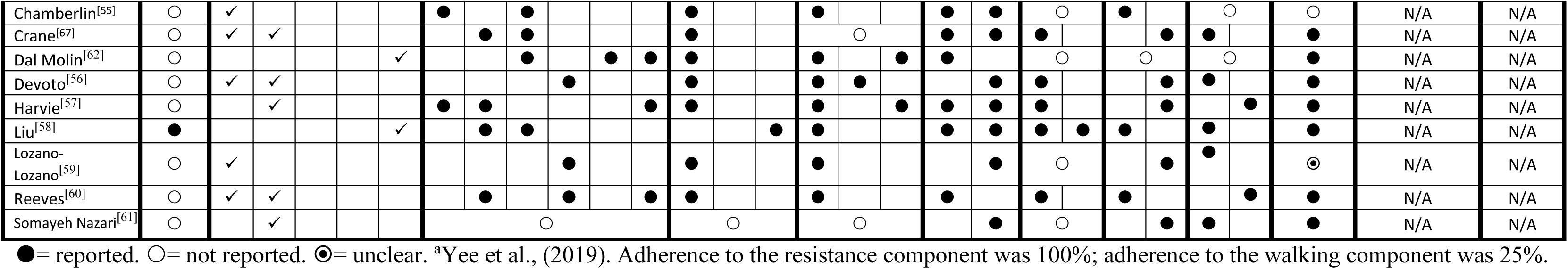
Summary of interventions according to the TIDieR checklist.

Six interventions involved one type of PA^[40,41,44–46,51]^ and six involved two^[39,42,43,47–49]^. The most commonly studied type of PA was aerobic^[41,46,47,49–51]^, followed by flexibility, stretching and mobility^[39,40,42,43,48]^, and balance/functional activity^[39,42,43,48,50]^. Only three full reports evaluated an intervention involving resistance/strength exercises^[47,49,50]^, and two reports of the same intervention evaluated a virtual assistant^[44,45]^. Most interventions used a combination of materials; five used videos^[39,40,43–45]^, five used handouts/information materials^[39,43–45,47]^, six provided some form of gym equipment or membership^[41,45–47,49,50]^. Six interventions used a wearable device^[41,45,47,49–51]^, and three provided nutrition support in addition to PA^[44,46,47]^.

Eight interventions were delivered by an exercise or healthcare professional, including; yoga instructors^[39,43,48]^, trained exercise specialists/physiologists^[41,47,49,50]^, and an experienced medical Qigong instructor^[42]^. Four interventions were delivered by a member of the research team^[44–46,51]^, and two used virtual assistants^[44,45]^. Eight interventions were delivered individually^[40,44,45,47–51]^, three via group sessions^[39,42,43]^, and one as a mixture of individual sessions and group cooking classes^[47]^. Seven interventions explicitly mentioned some element of home practice^[39–43,49,50]^. Four interventions were delivered exclusively remotely^[40,44,45,51]^, five in person^[39,42,43,46,48]^, and four were a mixture of in person and remote^[41,47,49,50]^.

Four articles did not report the duration of sessions^[41,46–48]^, and two were unclear^[44,45]^. Where duration was reported, two interventions involved sessions of >60 minutes^[39,43]^, and five involved sessions ≤60minutes^[40,42,49–51]^. Six interventions had sessions occurring weekly or less frequently^[39,41–44,47]^, and seven lasted for ≤12 weeks^[39,42,43,46–49]^. Tailoring was reported in eight interventions, often described as individualising the exercise based on performance, strength, progression, symptoms and metastases location^[41,44–47,49–51]^.

Medium to high (≥50%) adherence was reported in most studies (n=10)^[40,42–44,46–51]^, but was unclear in two^[39,41]^. In one study, adherence to 16 supervised sessions involving a brisk walk and resistance training was 100%, but only 25% of women adhered to the accompanying, unsupervised, walking component^49^. Attrition was varied, with two studies reporting low^[49,51]^, five medium (13 to 26%)^[40,43–45,50]^, and three reporting high levels of attrition (>26%)^[41,42,46]^. The highest attrition rate was 37%, from a study evaluating a Medical Qigong intervention^[42]^.

#### Study findings

Increases in time spent exercising were reported in three studies^[41,47,51]^, with one being statistically significant^[47]^, despite none being powered to detect a significant effect on these outcomes. Significant improvements were reported for handgrip strength, physical fitness, and additional physical assessments (e.g. 6-minute walk test, leg strength) across three studies of varying size (n’s=14, 40, 357)^[47,49,50]^. All three studies included both aerobic and resistance training exercises, with some supervision. Five studies reported on sleep outcomes^[43–46,51]^, with none reporting significant between-group differences, despite some trends towards improvement^[51]^. All were underpowered to detect effects on these outcomes.

Evidence for interventions improving fatigue was mixed: four studies, testing yoga^[48]^, seated exercise^[40]^, supervised resistance training plus unsupervised walking^[49]^, and a large scale (n=357) 9-month aerobic, resistance and balance programme^[50]^, found statistically significant improvements. A further three trials reported non-significant trends and/or improvements pre-post intervention, but not between groups^[42,47,51]^. The largest trial (n=357) found significant improvements in fatigue at 3-, 6-, and 9-months post-randomisation (Effect sizes = 0.14 to 0.24)^[50]^. This trial also reported significant improvements in health-related QoL and physical fitness at all follow-ups, alongside improvements in physical functioning at 6-and 9-months. Two studies, testing aerobic exercise^[41]^, and an in-home mHealth intervention^[45]^, did not improve fatigue.

Health Related Quality of Life, measured by the EORTC-QLQ-C30 or FACT-B, was significantly improved in two studies, involving both aerobic and resistance training^[47,50]^. Non-significant trends were reported in a further three (involving aerobic only/and resistance training) on global health scores and various subscales^[41,49,51]^. However three studies, involving Qigong^[42]^, cycling^[46]^, and a virtual assistant^[44]^, reported no significant differences on the FACT-B and/or SF36. One^[46]^ reported the attention control group (stretching exercises) significantly increased scores in the FACT-general score compared with those in the aerobic exercise group.

The majority (n=9) of studies reported no safety events and/or additional healthcare visits related to the intervention^[40–45,47,49,51]^, and two did not report on safety^[39,48]^. One pilot study reported that 36% of participants in the aerobic exercise intervention arm stopped a session early due to non-serious health events^[46]^, 73% (n=24) experienced at least one non-serious adverse event (AE) (compared to 7 AEs occurring in the attention control (stretching) group), and zero serious adverse events (SAEs) were observed. One phase III trial testing resistance training, aerobic exercise, and balance reported two exercise-related SAEs (a wrist fracture and sacral stress fracture), and 80 AEs requiring modifications to the exercise programme^[50]^.

Four studies included clinical outcome measures (e.g. inflammatory biomarkers), however these were not all reported within the included articles^[40,42,46,47]^. Of those that were, one reported no difference in tumour response to treatment between groups^[40]^, another reported no differences in haematological profiles between groups^[46]^, and one subgroup analysis (n=12) reported a non-significant increase in respiratory capacity in intervention participants^[47]^.

### Protocols and study registries

#### Characteristics of studies

We identified 15 ongoing trials. Most are being conducted in the USA(3)^[56,57,65]^, Europe(5)^[52,55,59,61,66]^, and Australia(4)^[53,58,60,62]^. Studies are also based in Iran^[63]^, Brazil^[64]^, and Canada^[54]^. Four studies are phase III RCTs^[52,55,62,65]^, seven are phase II or pilot/feasibility studies^[53,54,56,57,59,60,66]^ and the remainder did not describe the phase^[58,61,63,64]^. The planned or actual sample sizes range from 4^[57]^ to 260^[65]^ participants. Six studies will include individuals with an ECOG performance status of ≤2^[52,54,56,58,60,62]^, four studies with an ECOG of ≤1^[55,59,61,66]^, and one with a Karnofsky performance status of ≥80%^[57]^. The primary outcomes will include self-reported measures of fatigue^[52,53,55,58,65]^ and quality of life^[52,61,64]^, and self-reported^[57,60,62]^ and objective^[56,57,60,62]^ assessments of physical functioning/activity. Six studies are using co-primary outcomes; one combining self-reported fatigue with either time to deterioration of health status or HrQoL^[52]^, one assessing self-reported physical function and whole body lean mass^[62]^, one assessing objective functional capacity alongside self-reported QoL^[61]^, and two using objective and self-reported assessments of physical functioning/activity^[57,60]^. Other primary outcomes include progression free survival^[59]^, liver enzymes^[63]^, change in lesion size^[54]^, feasibility/acceptability^[53,66]^, and safety^[53]^. Follow up periods range from 8 weeks^[63]^ to 2 years^[54]^.

Three studies are using usual care as a comparator^[53,57,61]^, six are using active controls^[55,58–60,64,65]^. Four^[52,54,56,62]^ are using varying approaches to enhanced usual care e.g. a leaflet^[56]^ or PA recommendation^[52]^.

#### Intervention characteristics according to the TIDieR checklist

Table 3 summarizes the intervention characteristics for trial registries and protocols, according to the TIDieR checklist, with full data extraction notes in Appendix 3. One report states topics of discussion in behavioural counselling will be guided by the transtheoretical model of behaviour change^[60]^. No others describe a theoretical basis to intervention development. Eight interventions are incorporating one type of PA^[52,54,57,59–61,63,64]^, and four are incorporating two types^[55,58,62,65]^. Aerobic exercise is the most common (n=10)^[52–54,56–58,61,62,65,66]^, followed by strength/resistance training (n=8)^[53,56,58,59,62,63,65,66]^. Six interventions are incorporating wearable devices^[52,56,57,60,64,65]^, five include access to gym equipment or a membership^[53,54,58,61,62]^, and two involve an app^[52,64]^.

Most interventions (n=13) are being delivered by exercise professionals or healthcare professionals; including physiotherapists/exercise professionals^[52–54,56,58,59,62,64,66]^, dieticians/nutritionists^[54,59,62,64]^, nurses^[57,64]^, psychologists^[64]^, a multidisciplinary team^[61]^, a trained health coach^[65]^, and a certified eurythmy therapist^[55]^. One intervention is being delivered by a doctoral student^[60]^.

Nine interventions are individual^[52–54,57,59–62,64]^, one involves small groups of 1-4 people^[55]^, and one includes either individual or group sessions of up to four people^[58]^. Five interventions are encouraging home practice^[52,53,55,59,64]^. Four interventions are remote^[52,56,62,64]^, six are in person^[53,55,58,61,63,66]^, two are using a combination of in person and remote sessions ^[54,60]^, and three are offering a choice^[57,59,65]^.

Most interventions include sessions >60 minutes (n=10)^[52–56,58,59,60,62,65]^, with one including longer sessions (90-120 minutes) for the first and final session^[60]^. Five interventions have sessions occurring weekly or less frequently^[54,55,57,60,62]^, while sessions are held more than weekly for nine interventions^[52,53,56,58,59,61,63,65,66]^, taking place up to three times a week^[53,56,63,65,66]^. The duration of interventions ranges from 6 weeks^[58]^ to 12 months^[62]^.

Most interventions report tailoring; five describe tailoring based on a combination of factors including metabolic equivalent time minutes, treatment regime, current activity and energy levels, heart rate and performance^[52–54,59,62]^. One focuses tailoring on heart rate, rate of perceived exertion and responses during the session^[56]^, five describe tailoring based on individual capabilities and needs^[55,58,60,63,65]^, one describes providing personalised advice in a chat^[64]^, and one states the intervention is individualised but does not specify how^[61]^. Two interventions do not report any tailoring^[57,66]^.

## Discussion

This systematic review demonstrates PA interventions in people with advanced and MBC are safe, well adhered to, and have the potential to improve fatigue, health-related QoL, physical fitness and physical functioning over the short and medium term. However, as many trials are still ongoing and most completed studies only tested for feasibility, evidence-based recommendations on frequency, duration, delivery mode, and intensity cannot yet be made.

Furthermore, trials with longer follow ups (>9 months) are needed to understand if activity levels and improved outcomes are sustained after intervention periods are completed. The overall methodological quality of the completed studies was moderate. While most employed appropriate study designs, outcome measures and randomisation procedures, common limitations included differential treatment between groups, inadequate reporting on allocation concealment of outcome assessors, and unclear reporting of group differences on follow-ups.

The type of interventions being evaluated vary from yoga, walking, and strength training, to in-home mHealth personal assistants. Interventions that included both a strength/resistance element and aerobic element seemed to be more effective at improving fatigue, QoL, and fitness than other activity types. Adherence levels were high overall, but dropped off over time and were generally lower for unsupervised components (mHealth^[44]^ and walking^[49]^).

The variety in activity type reflects the current lack of clinical guidance. Furthermore, a ‘one-size-fits-all’ approach is unlikely to be effective in this group, given the vast individual differences in physical capabilities, symptoms, and preferences^[30,67]^. Literature from other populations emphasizes enjoyment as key to adherence^[68,69]^, so patient preferences and individual barriers and facilitators to being active should be considered. While individual tailoring was evident in most studies, the specifics were not always reported. Information on the development of existing interventions is sparse, with only four stating some theoretical basis. Intervention development guidance advocates for theory-based interventions, to increase our understanding of the mechanisms of action^[70]^ and increase effectiveness: PA interventions using techniques linked to Control Theory^[71]^ are twice as effective as other interventions^[72]^. Future interventions should have a clearly described and reported theoretical basis.

The field of PA in MBC is growing, with 15 ongoing trials identified over the last 13 years. Within these ongoing trials there has been a greater emphasis on evaluating interventions which include strength/resistance training. This reflects the growing recognition of the importance of strength training for individuals with cancer, including breast cancer, to reduce the risk of cancer-related fatigue, muscle loss and impaired QoL^[14]^. Early evidence from a small number of completed studies in this review suggest interventions involving strength training components could also improve fatigue and QoL in MBC^[47,49,50]^.

### Strengths

This is the first systematic review looking at PA specifically in MBCs. All screening, data extraction and quality assessments were conducted in duplicate. Including protocols and ongoing trials ensured a comprehensive overview of the current research in this field, providing a single resource for researchers, clinicians, and affected individuals.

### Limitations

Non-English language publications were excluded. We did not extract information on behaviour change techniques (BCTs)^[73,74]^ as this was beyond the scope of this review, and most papers did not explicitly report them. Therefore, the granular level of intervention content from a behaviour change perspective is missing.

### Implications for clinical practice

Evidence to date suggests women with MBC should be encouraged to be active, and offered access to appropriate programmes that include both aerobic and strength/resistance-based exercises. Consideration of how to implement effective interventions within healthcare services is needed. Existing studies suggest that tailored interventions delivered by qualified exercise professionals can improve various outcomes, and are acceptable to those participating. However, evidence-based recommendations on specific frequency, duration, delivery mode, and intensity cannot yet be made. The accessibility of available PA programmes should be considered when making recommendations, to ensure potential inequalities are not increased; for example, highlighting online programmes for those in rural areas, or arranging for translators to overcome language barriers.

### Implications for future research

Fully-powered definitive trials, with cost-effectiveness evaluations, longer term (>9 months) follow-ups, and that properly evaluate all intervention components, are required. Interventions designed with patient and healthcare professionals’ involvement, which have a theoretical basis, are also needed. Most interventions were tested in parallel group designs or with crossover arms, which cannot provide evidence on whether all, or only some intervention components are effective. Future trials could consider novel trial designs (e.g. complex factorial trials) within the MOST framework to more efficiently design, optimise and evaluate multi-component interventions^[75]^. Consistency of outcome measures across trials would increase our understanding of the effectiveness of PA on difference outcomes. Future studies could include descriptions of the BCTs used, to enhance our knowledge of the active components of interventions^[76]^. Improved reporting of participant characteristics is needed to assess sample representativeness and whether certain groups are being excluded. Those that did report participant characteristics included samples that were not very diverse, with most participants being 50-60 years old and White.

### Conclusion

Evidence on the efficacy and safety of physical activity for women with MBC is growing. More trials are required before specific, evidence-based recommendations on PA type, duration, mode of delivery, and frequency can be made, along with their long-term effectiveness on clinical outcomes. However, existing studies provide initial evidence that exercise can be safe, well adhered to, and improve various outcomes including fatigue and QoL, for women with MBC. Physical activity should be actively encouraged and adequately supported by healthcare professionals involve in their care.

## Funding acknowledgements

This review was supported by funding from Yorkshire Cancer Research (L389SS, L389RB). This report is independent research supported by the National Institute for Health Research NIHR Advanced Fellowship, Prof Samuel Smith NIHR300588. The views expressed in this publication are those of the author(s) and not necessarily those of the NHS, the National Institute for Health and Social Care Research or the Department of Health and Social Care. The funders had no role in the design of the study, data collection, analysis, interpretation of data, and in the writing of this manuscript.

## Author contributions: CRediT

Conceptualisation: SGS, RJB

Data curation: LHH, SMCG, SH, AF, NK, RJB, SGS

Formal analysis: LHH, SMCG, SH, AF, NK, RJB, SGS

Funding acquisition: SGS, RJB

Investigation: LHH, SMCG, SH, AF, NK, RJB, SGS

Methodology: SGS, RJB, LHH, AF, NK

Project administration: LHH, SGS, RJB

Resources: N/A

Software: N/A

Supervision: SGS, RJB

Validation: LHH, SMCG, SH, AF, NK, RJB, SGS

Visualisation: SMCG, LHH

Writing – original draft: LHH, SMCG, RJB, SGS

Writing – review and editing: LHH, SMCG, SH, AF, NK, RJB, SGS

## Appendix 1. PRISMA 2020 Checklist

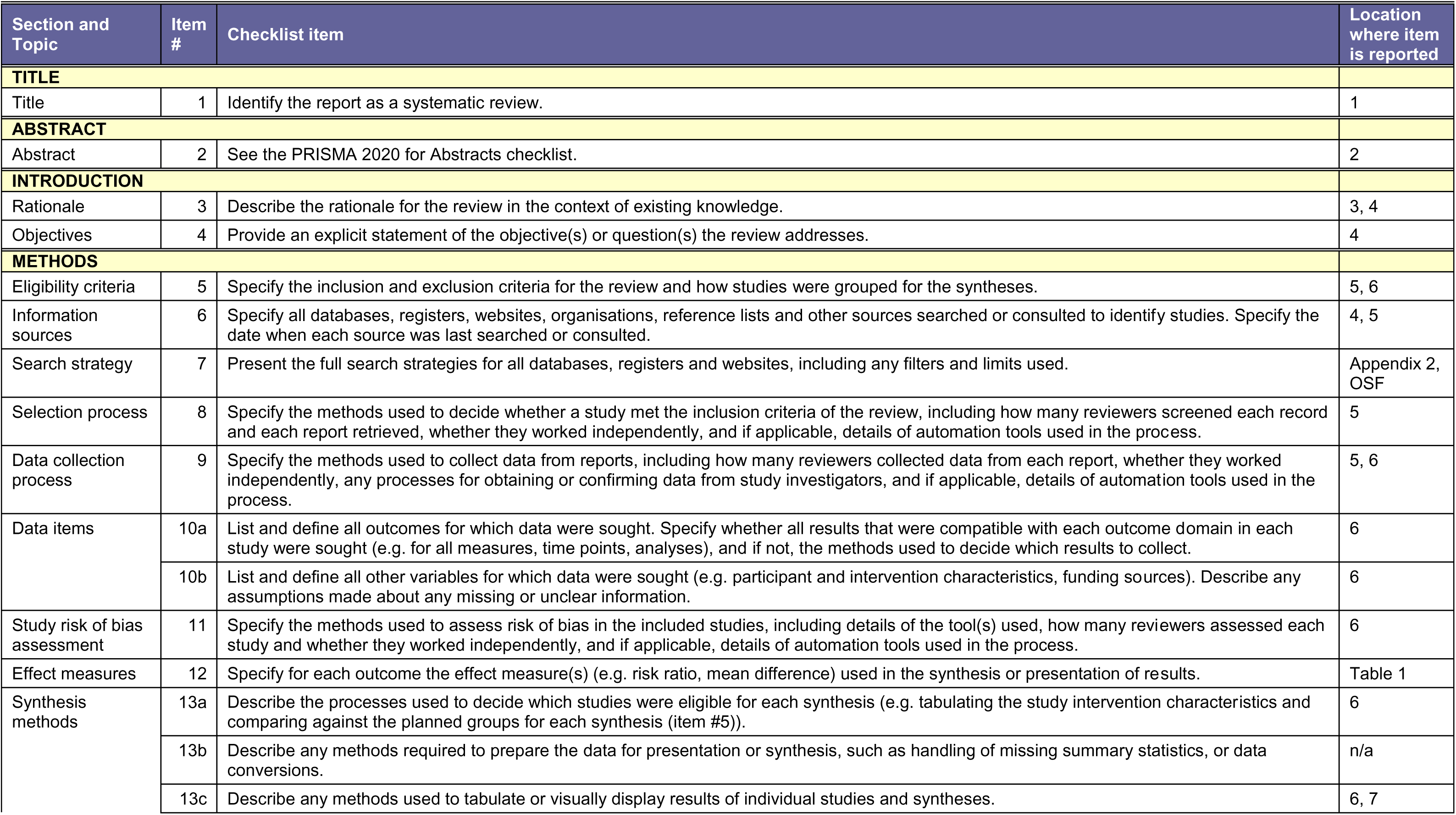
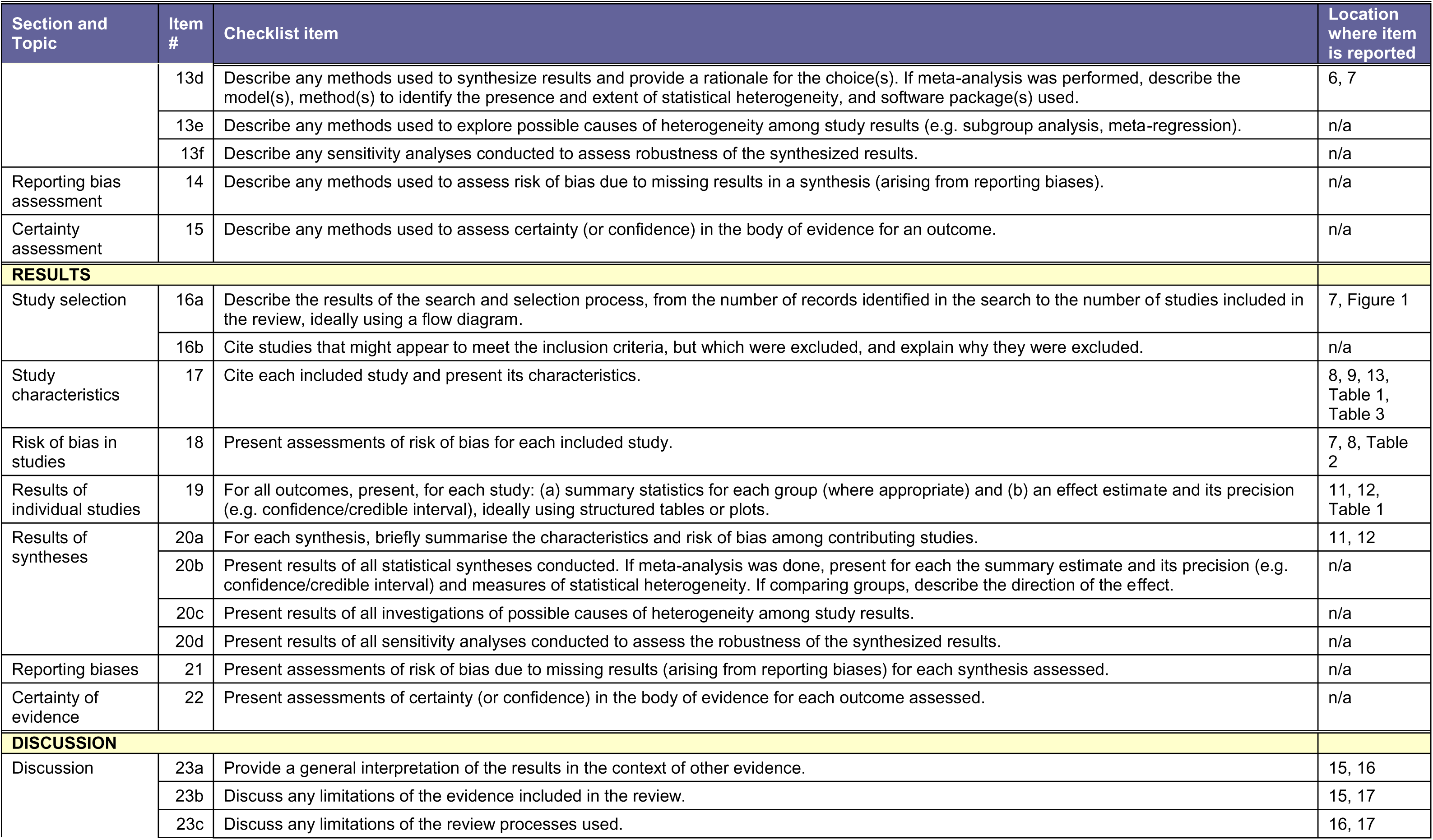
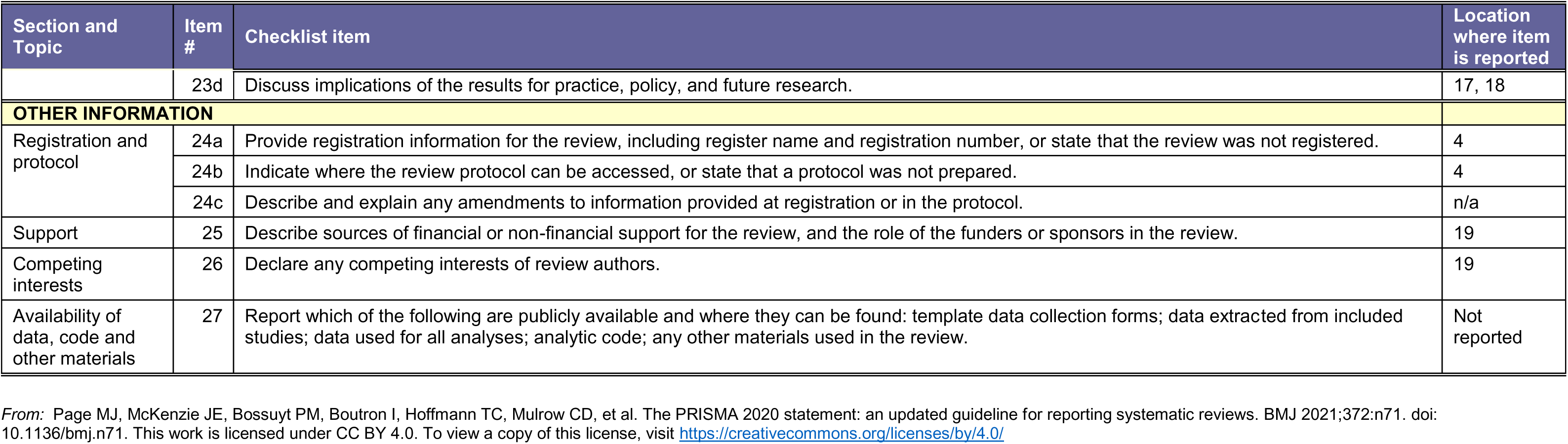

## Appendix 2. Search Strategy for Medline

1 exp Breast Neoplasms/ and exp Neoplasm Metastasis/
2 (metasta* adj4 (breast or mammary)).tw,kf.
3 ((advanced or “stage 4” or “stage IV”) adj2 (breast or mammary)).tw,kf.
4: 1 or 2 or 3
5: exp Exercise/
6 exp Sports/
7 exp Dancing/
8 (physical adj3 activit*).tw,kf.
9 (exercise or exercises or exercising).tw,kf.
10 (physical adj3 (therap* or train* or conditioning)).tw,kf.
11 ((muscle or strength* or resistance or circuit* or interval or weight*) adj3 train*).tw,kf.
12 exp Swimming/
13 exp Walking/67787
14 exp Running/
15 exp Jogging/
16 sport*.tw,kf.
17 exp Fitness Centers/
18 gym*.tw,kf.
19 aerobics.tw,kf.
20: 5 or 6 or 7 or 8 or 9 or 10 or 11 or 12 or 13 or 14 or 15 or 16 or 17 or 18
21 controlled clinical trial.pt.
22 randomized controlled trial.pt.
23 randomized.ab.
24 placebo.ab.
25 clinical trials as topic.sh.
26 randomly.ab.
27 trial.ti.
28: 21 or 22 or 23 or 24 or 25 or 26 or 27
29 exp animals/ not humans.sh.
30: 28 not 29
31: 4 and 20
32: 30 and 31

## Appendix 3. Data extraction for interventions according to the TIDieR checklist

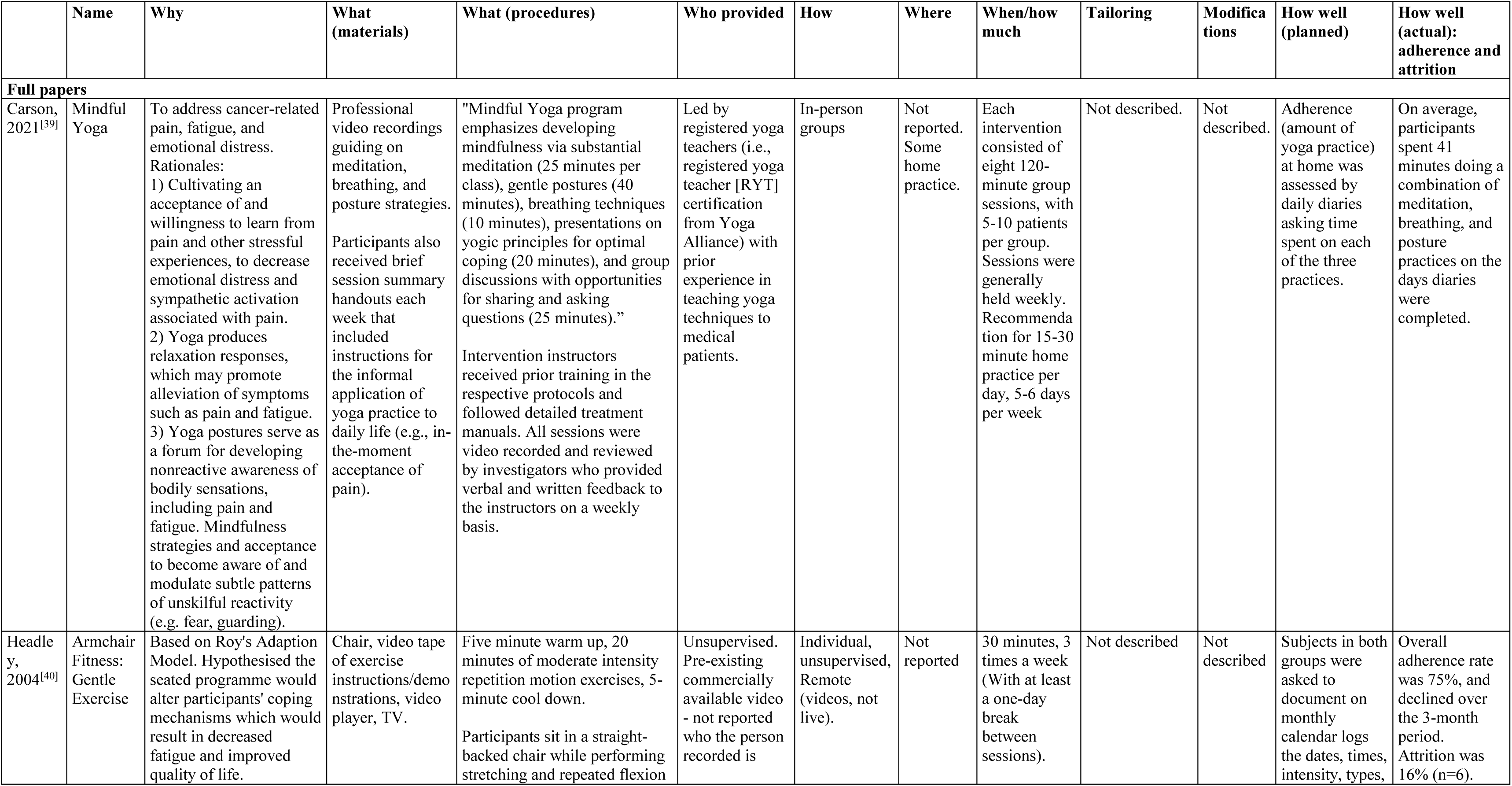
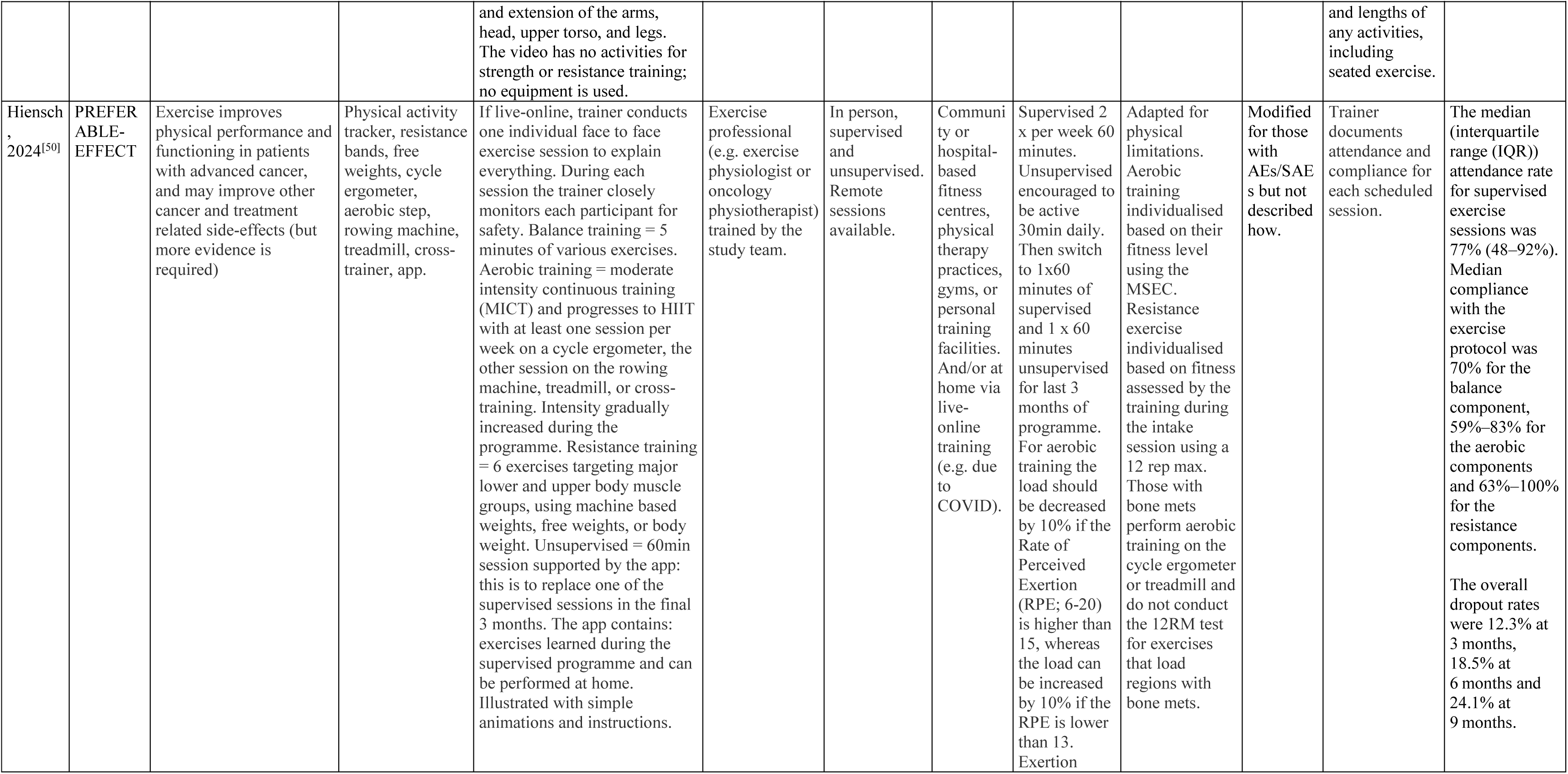
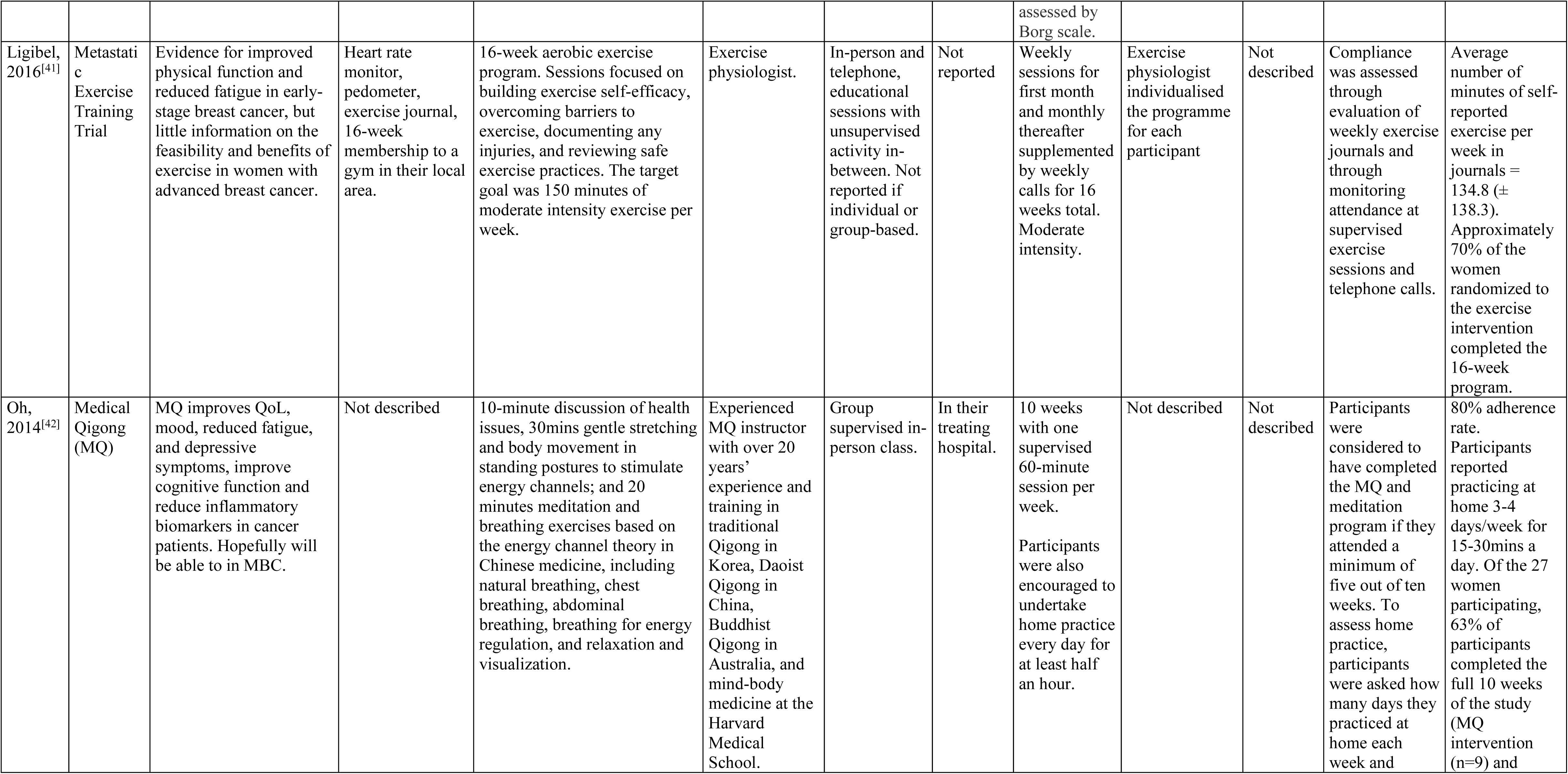
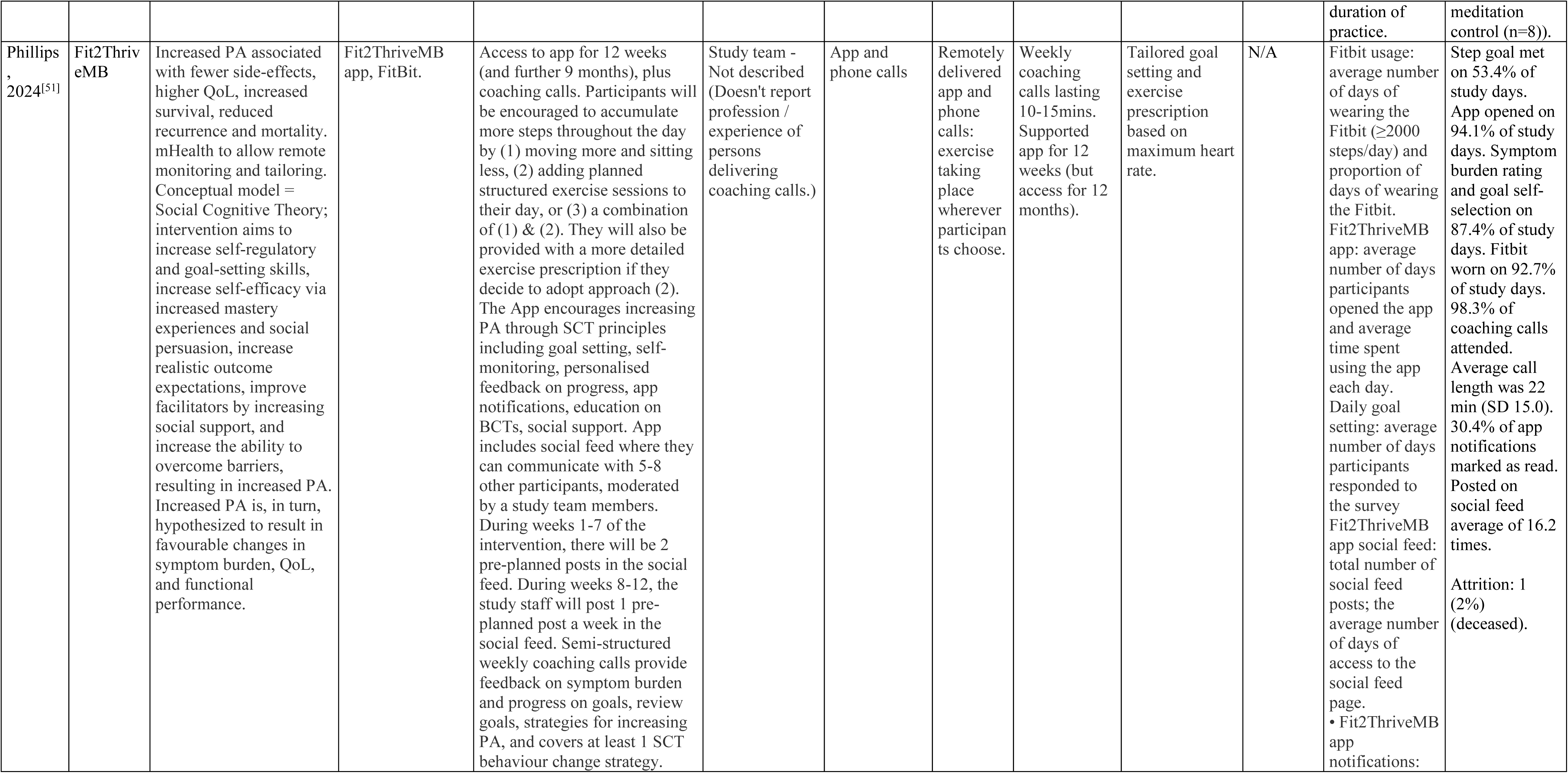
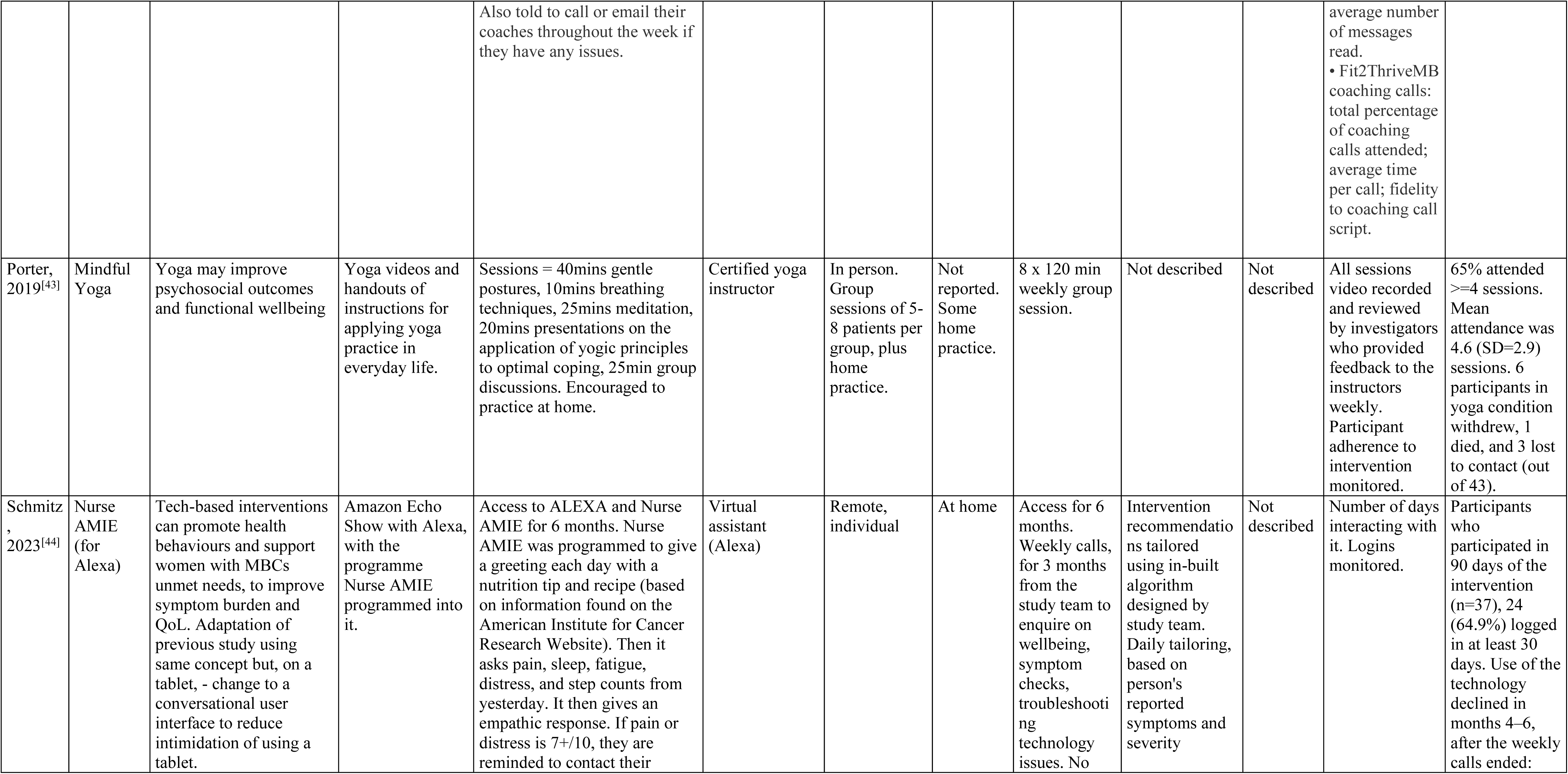
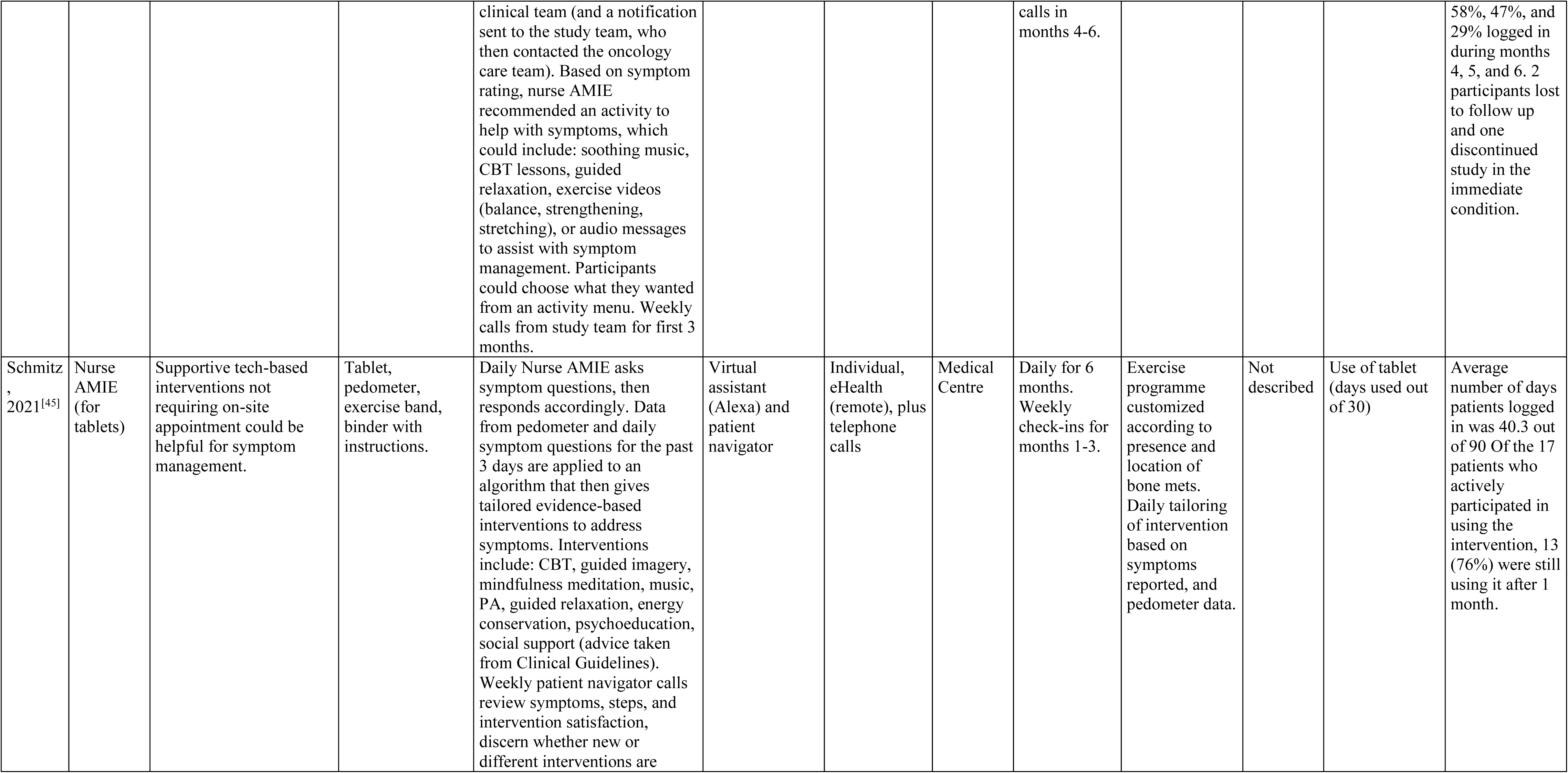
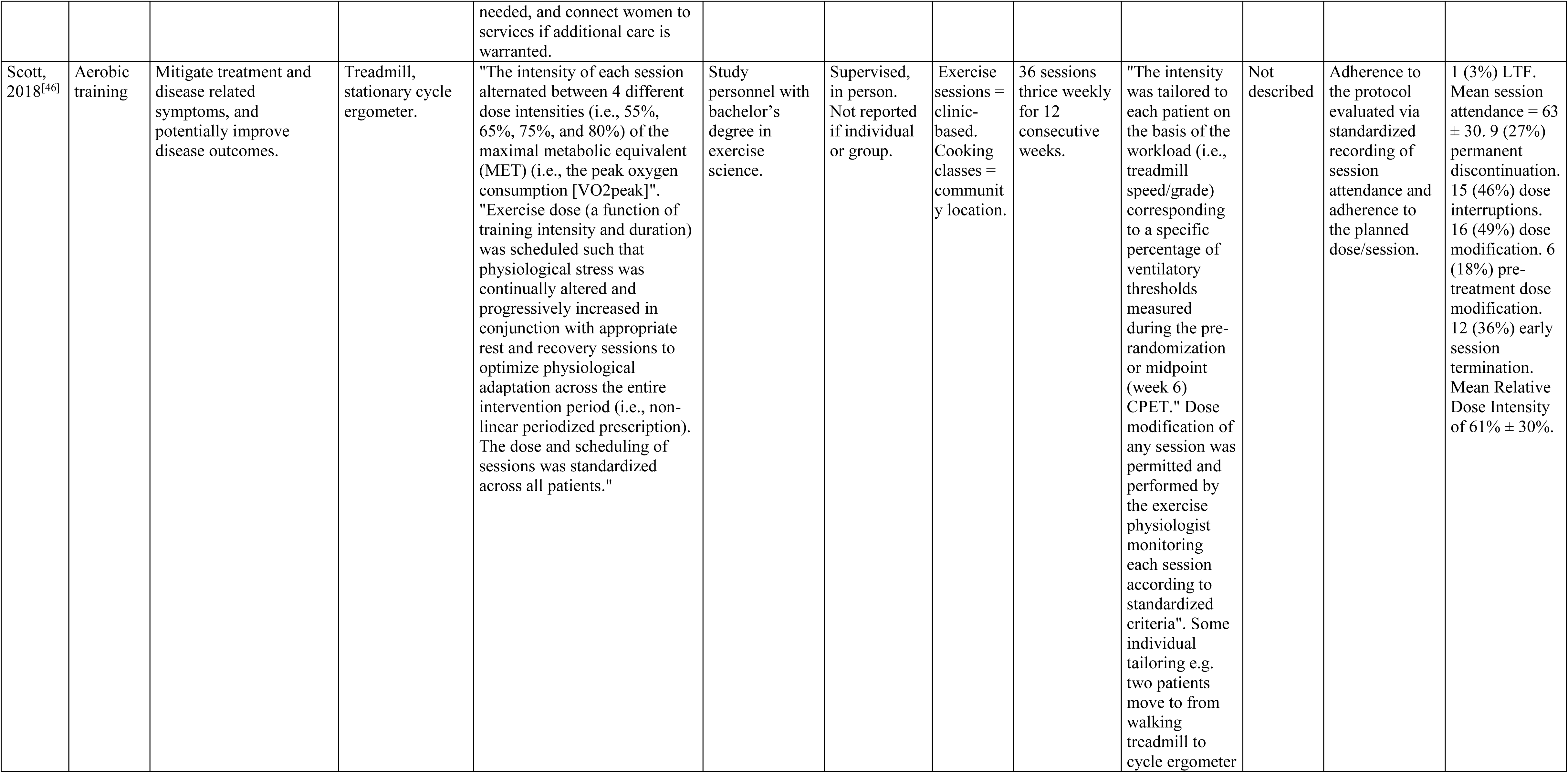
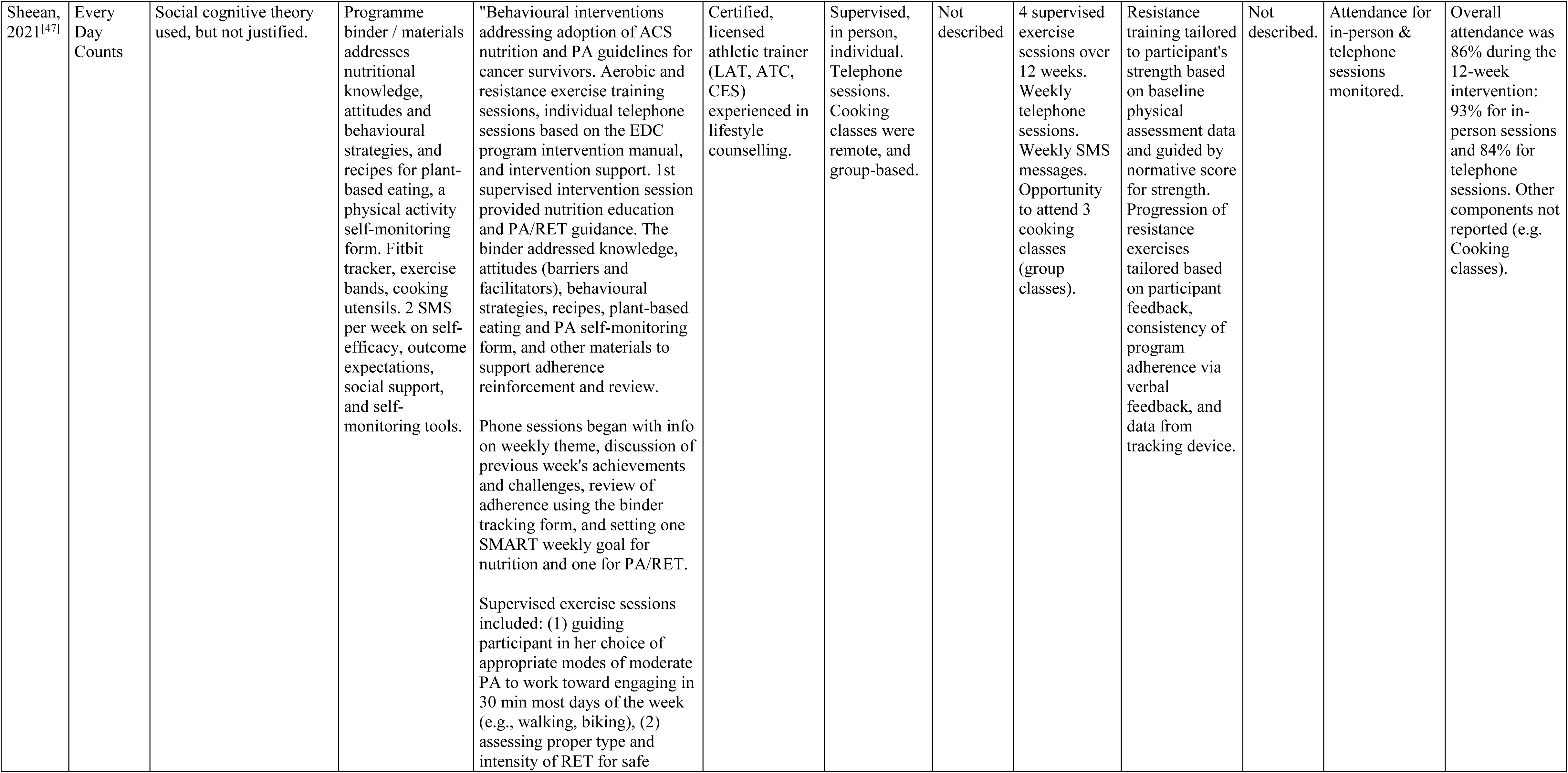
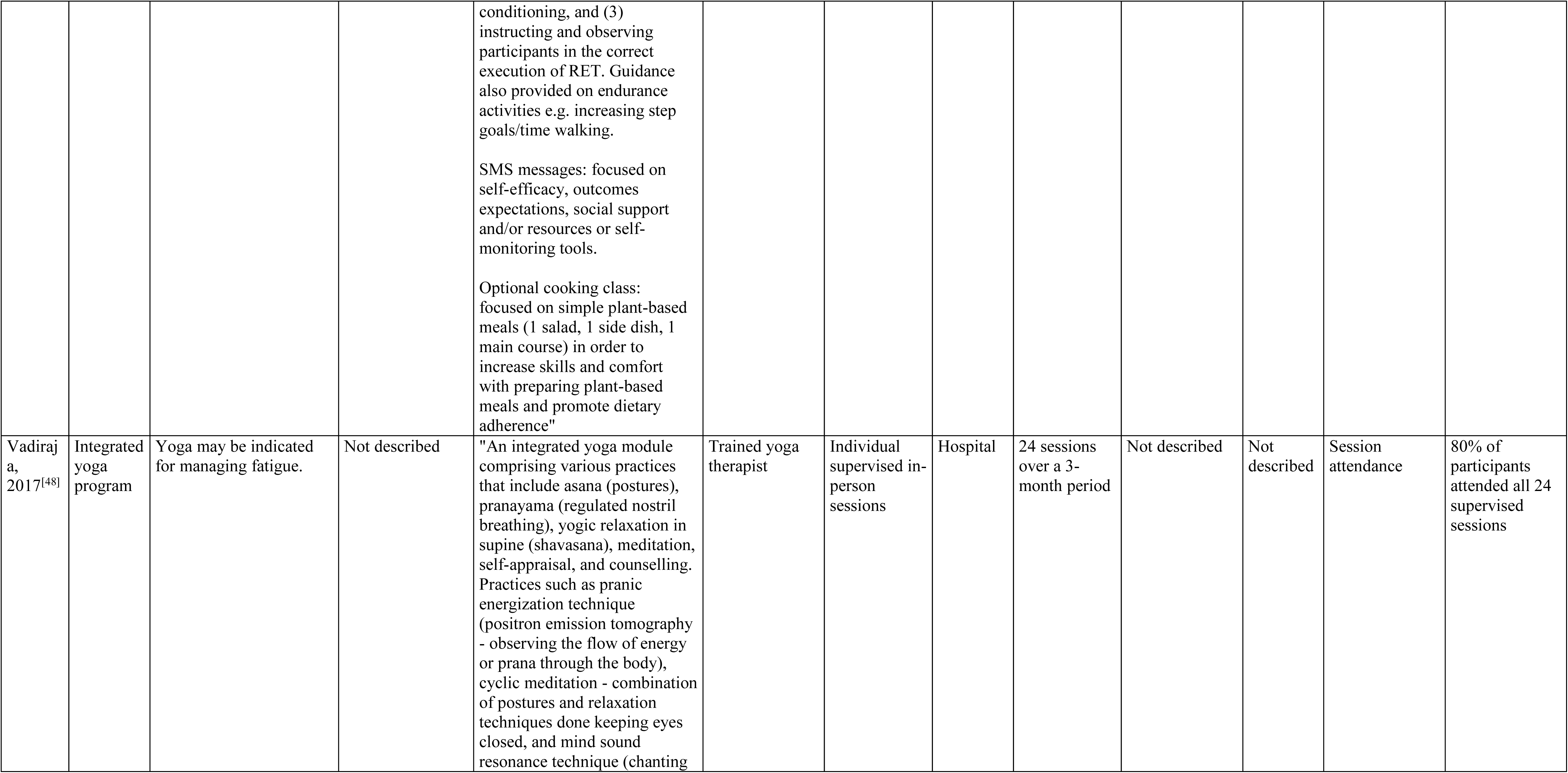
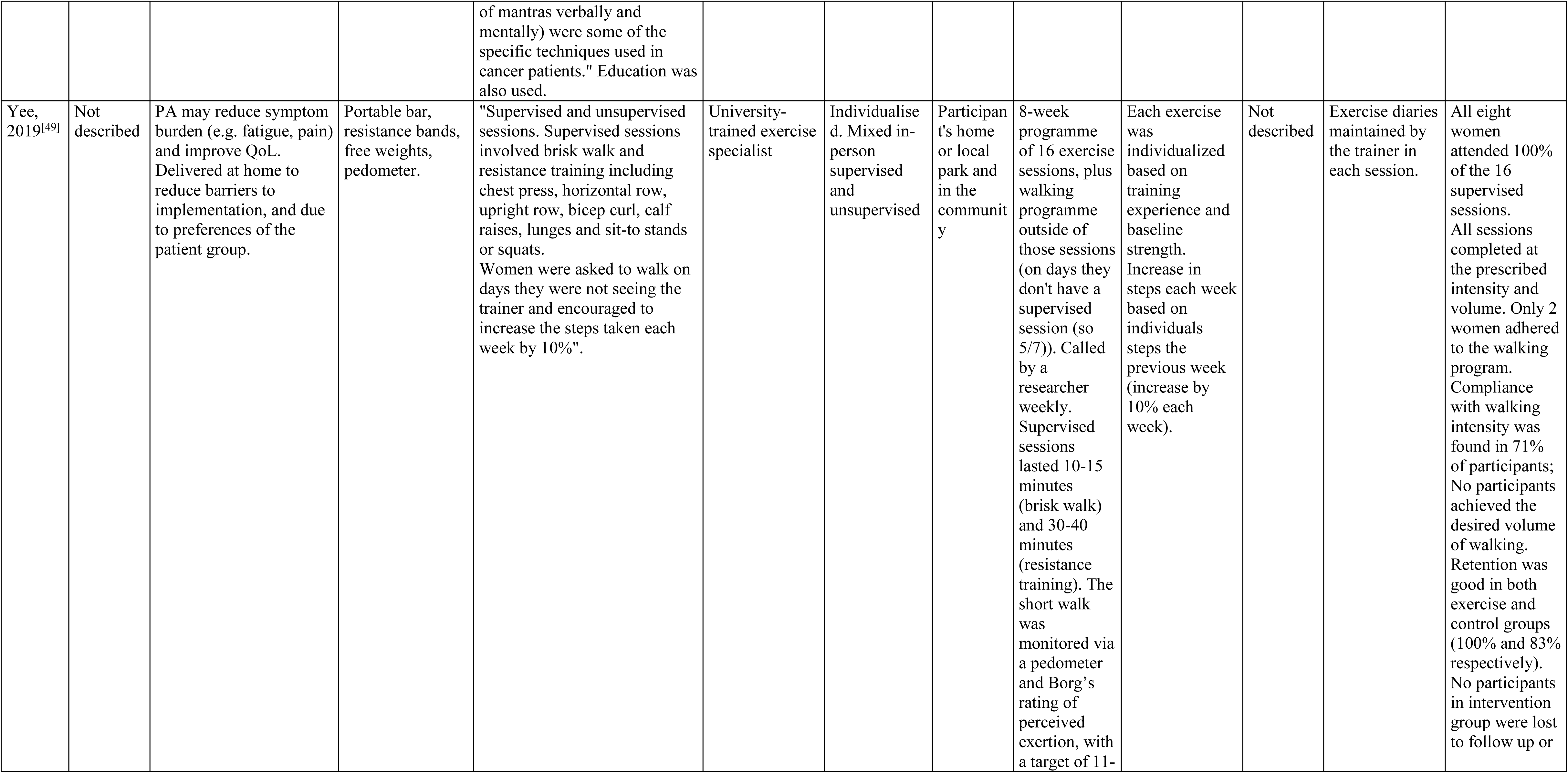
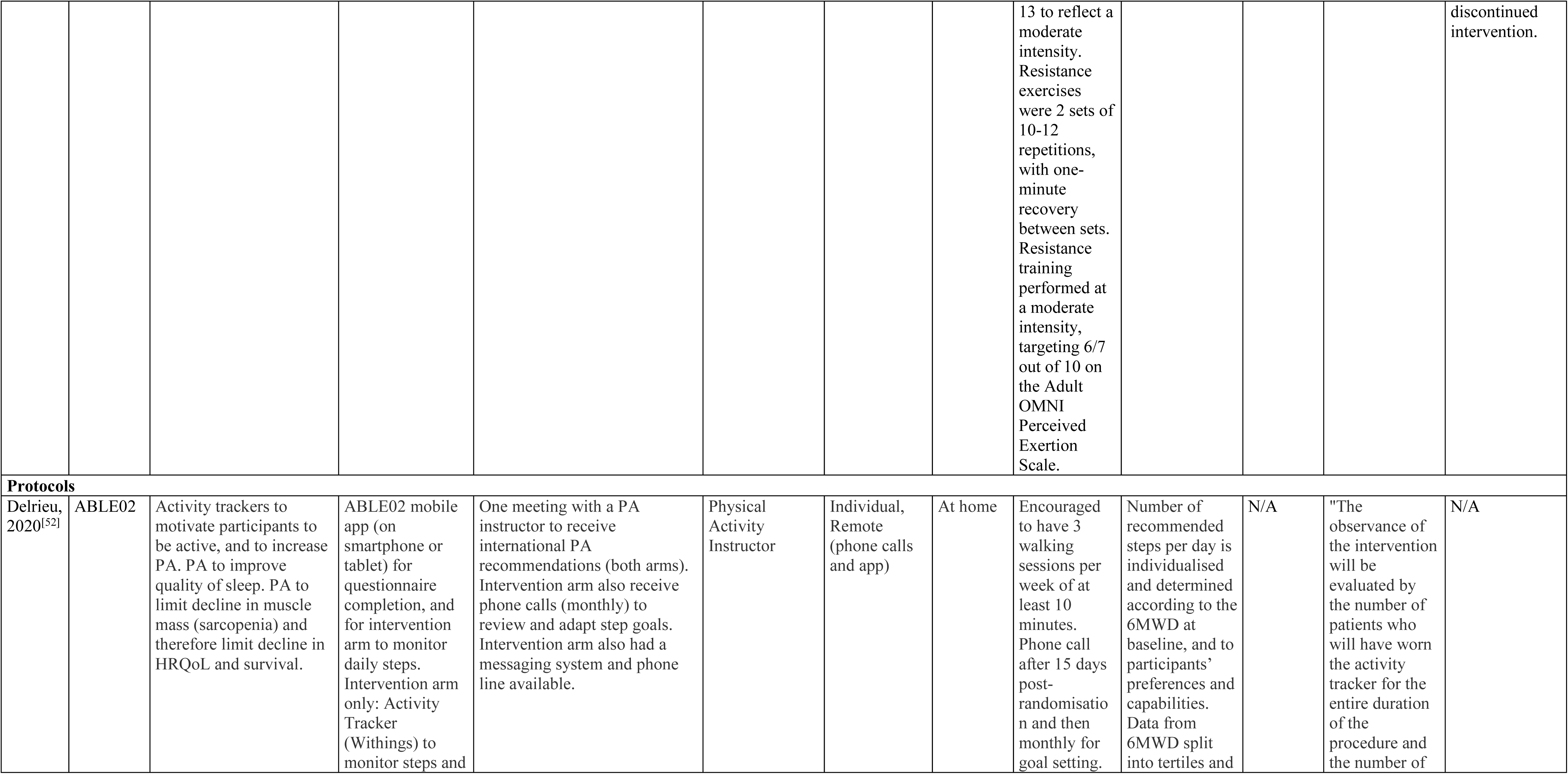
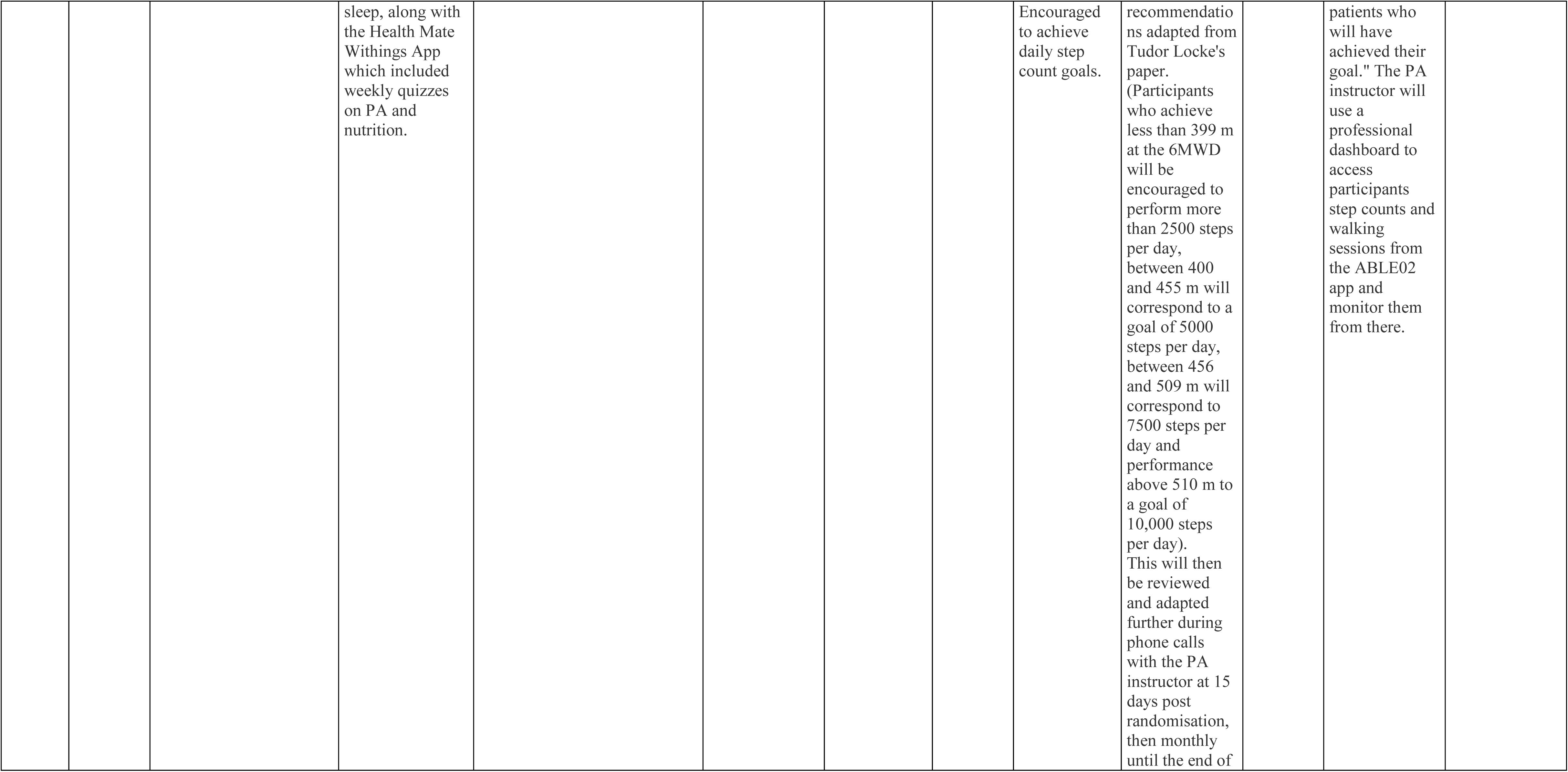
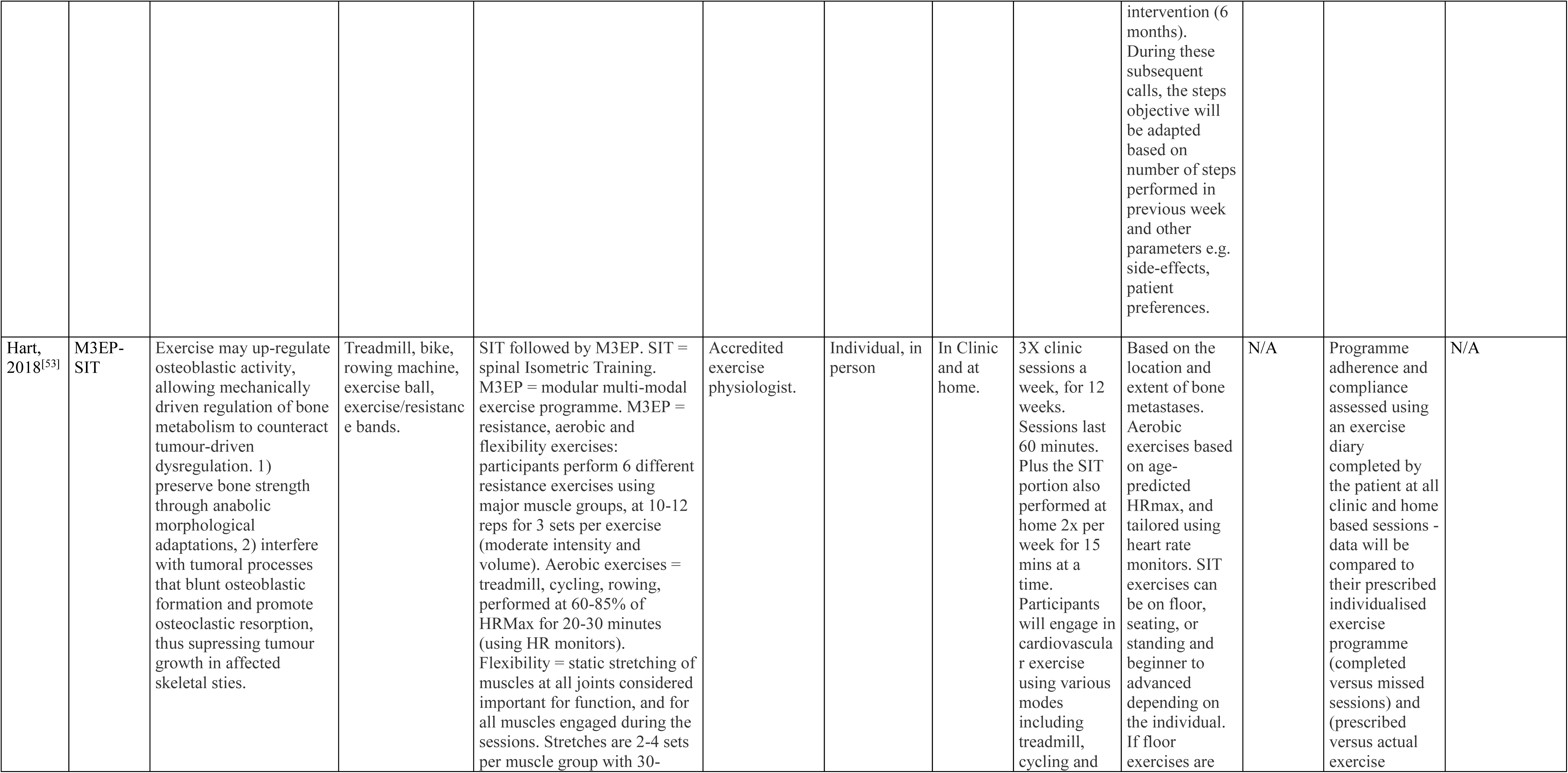
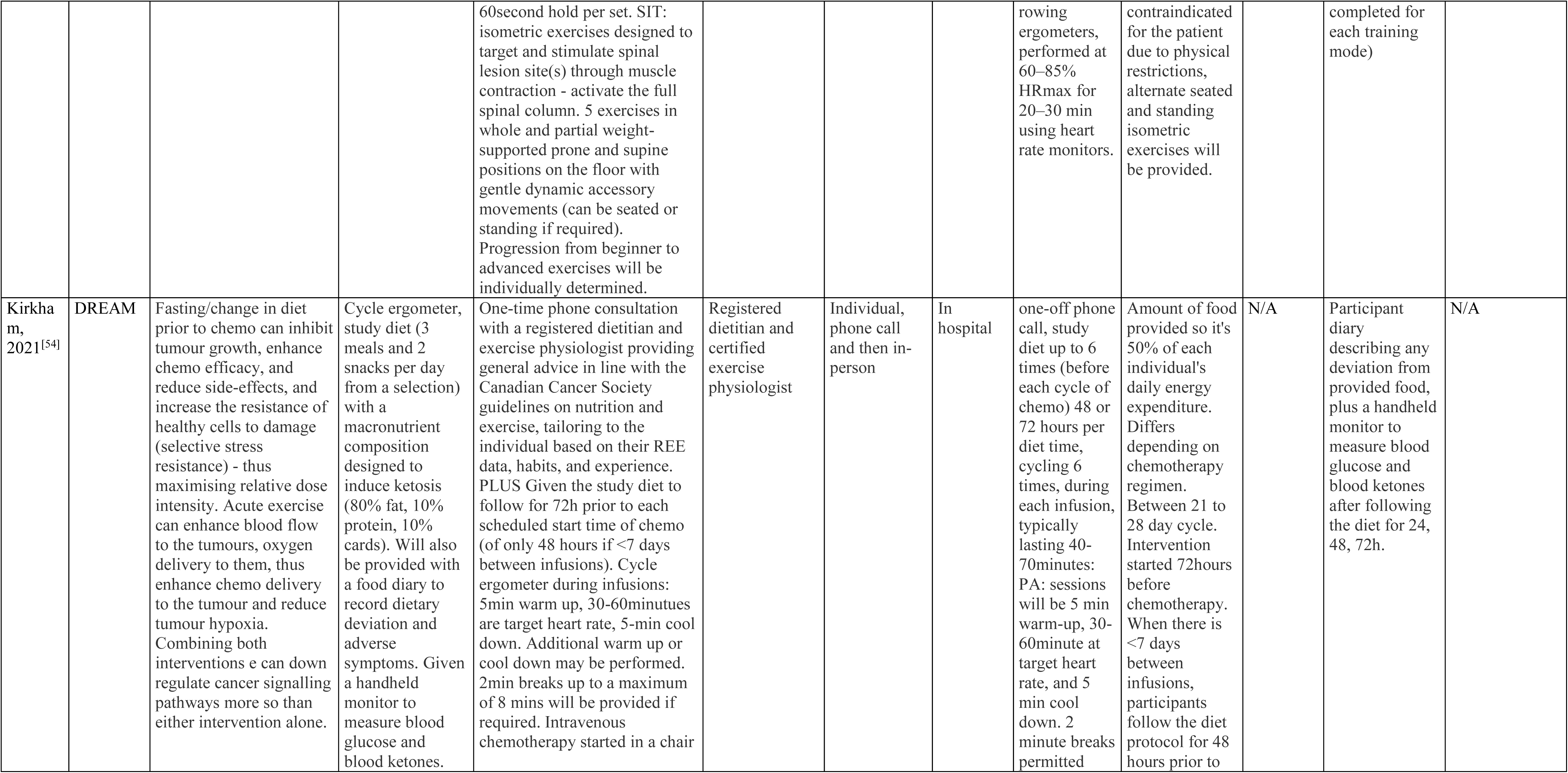
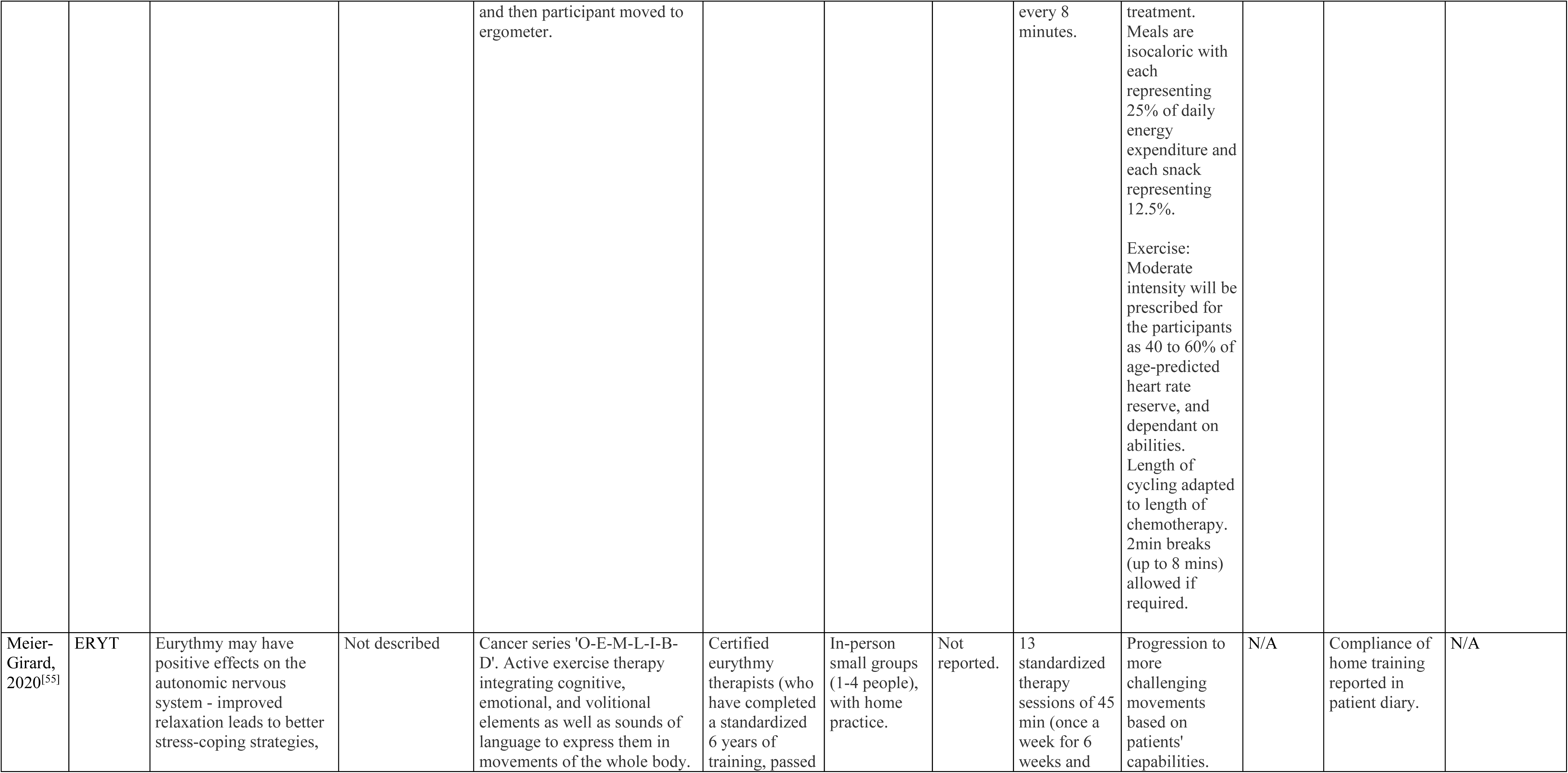
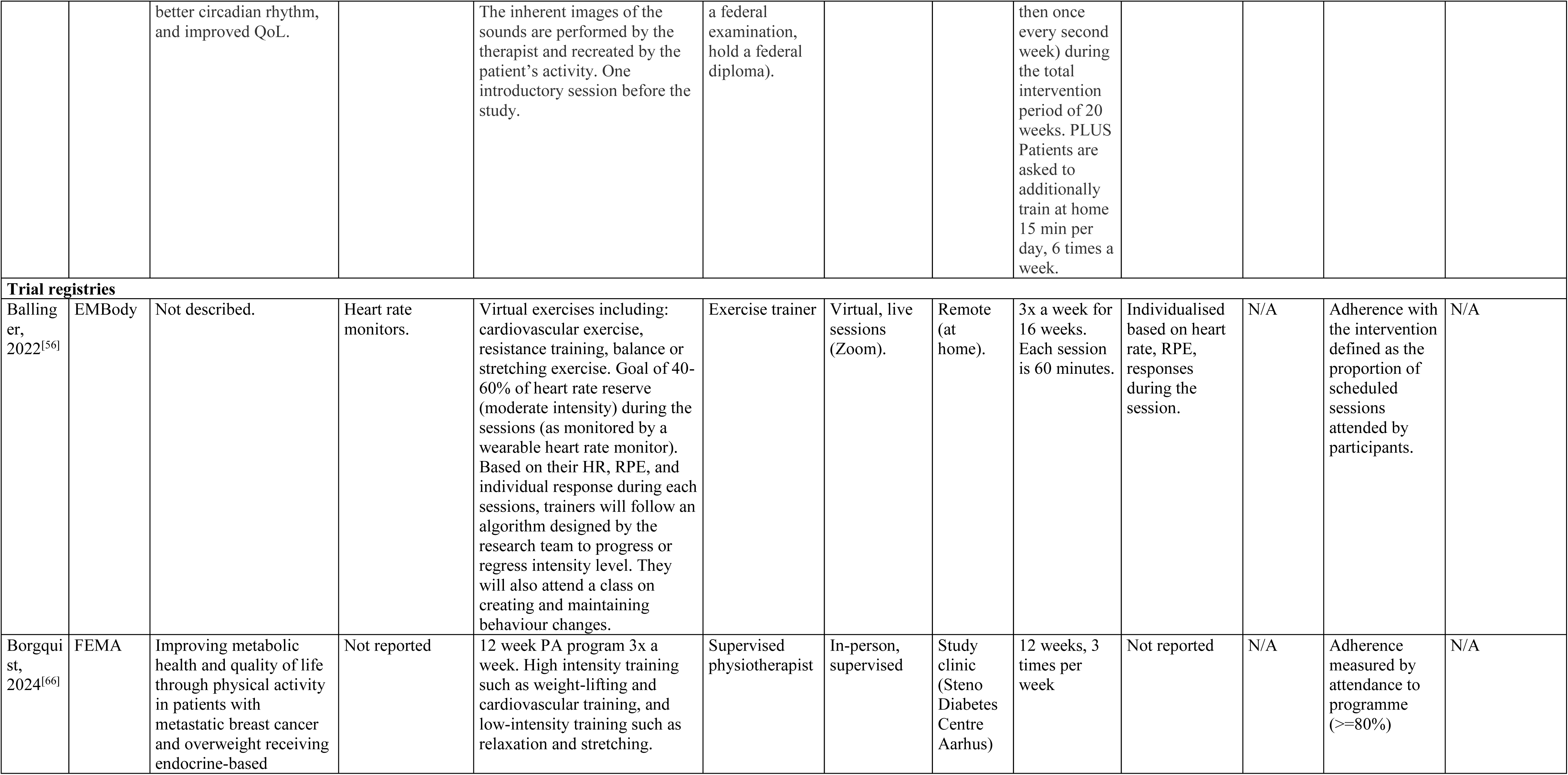
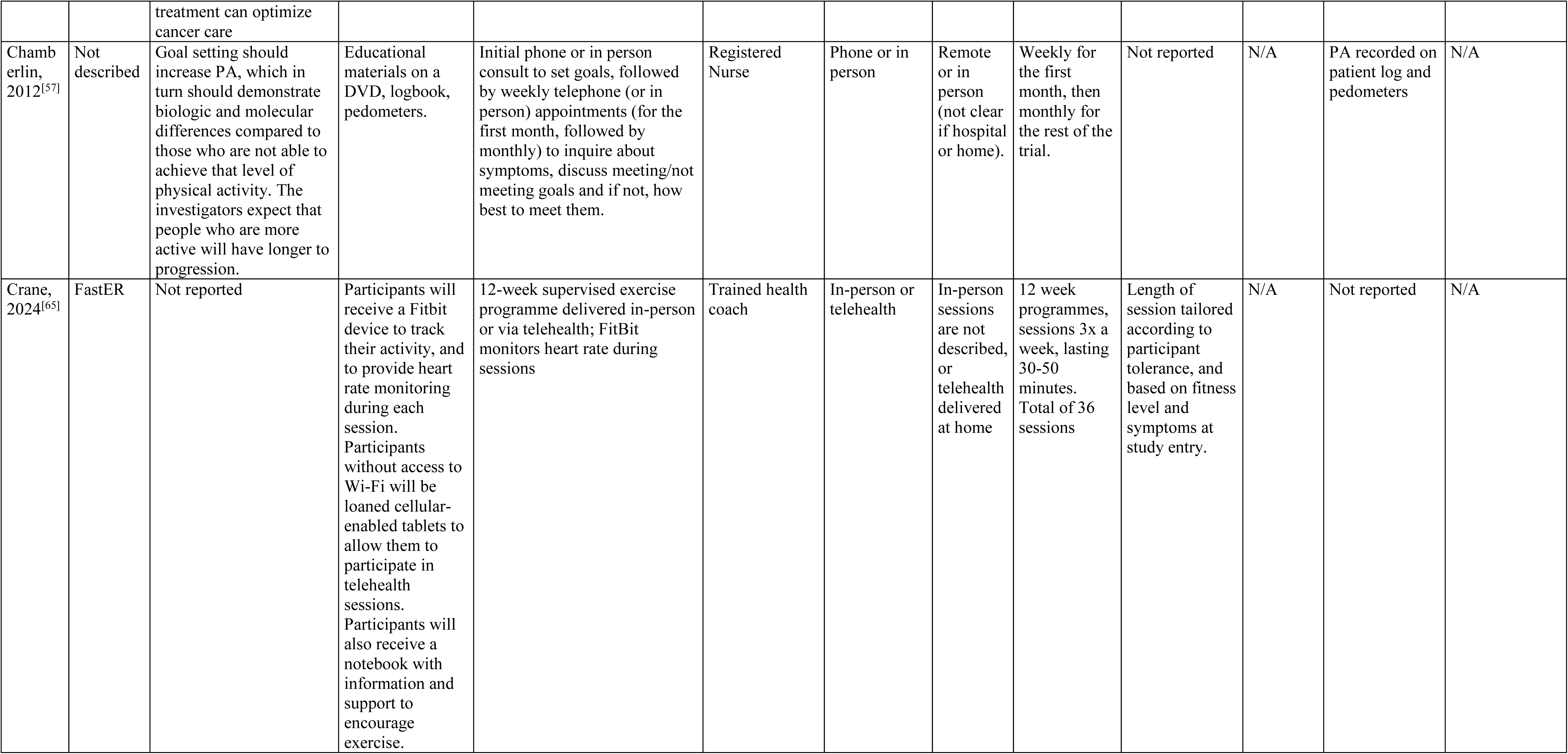
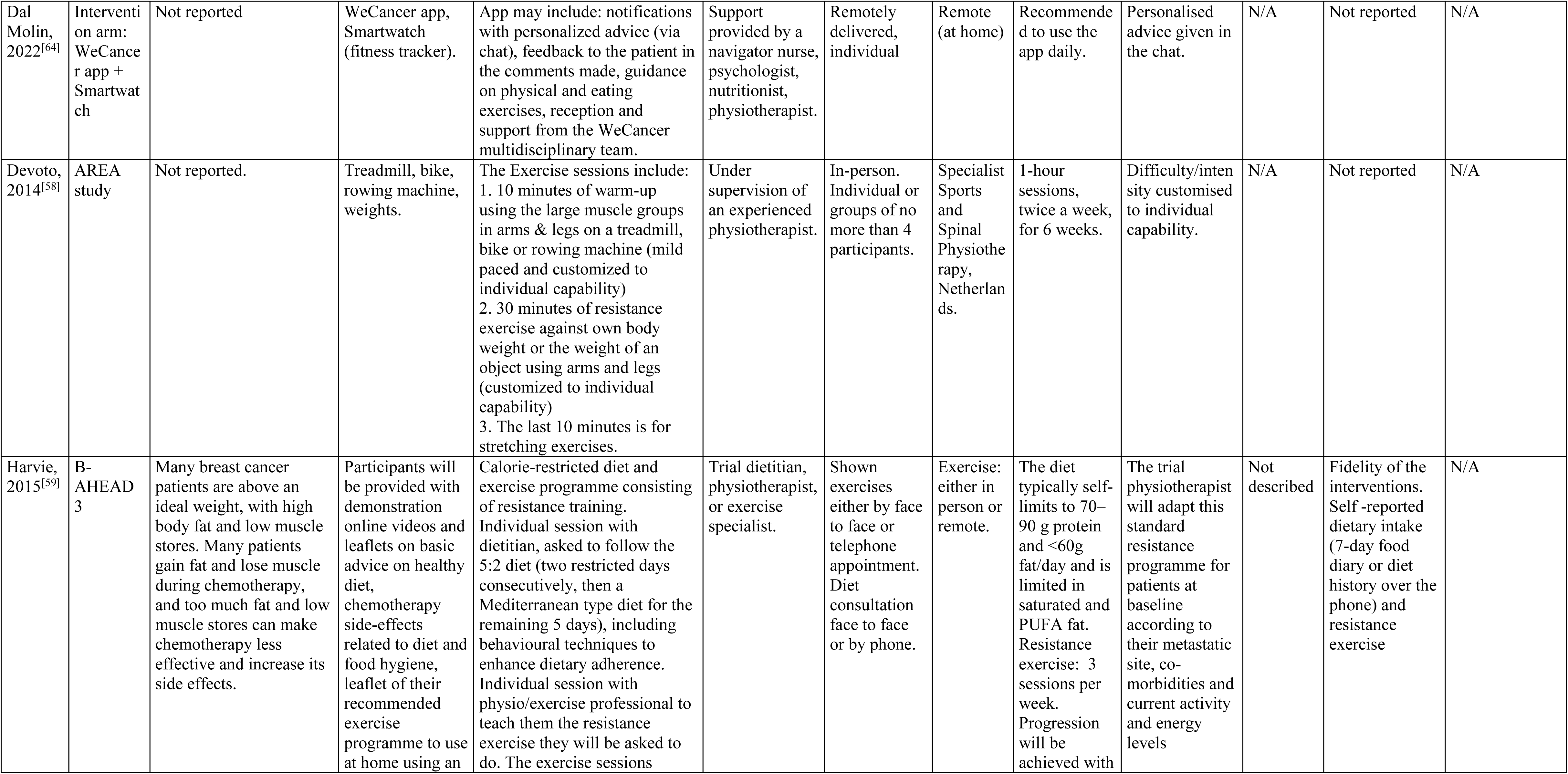
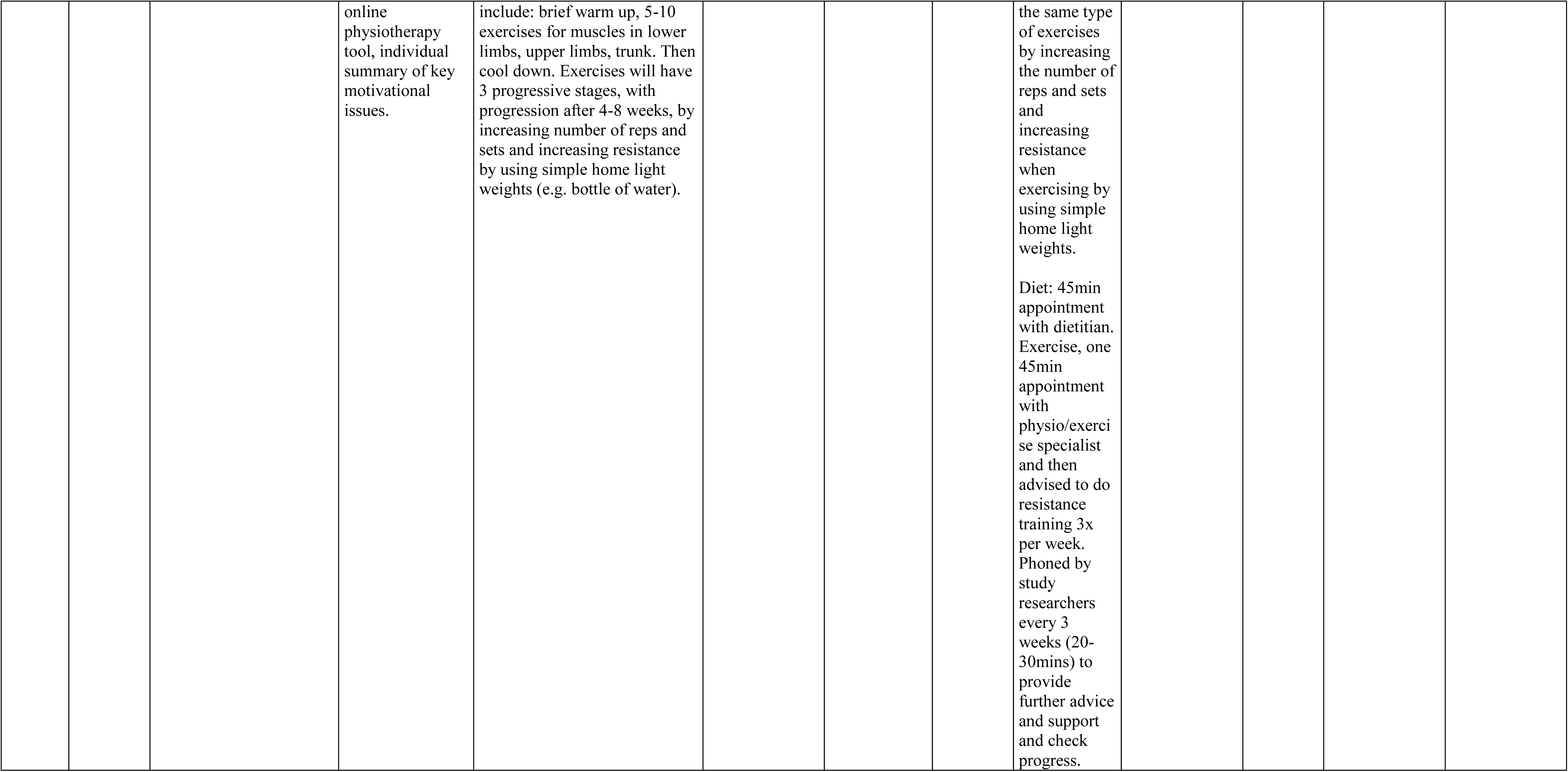
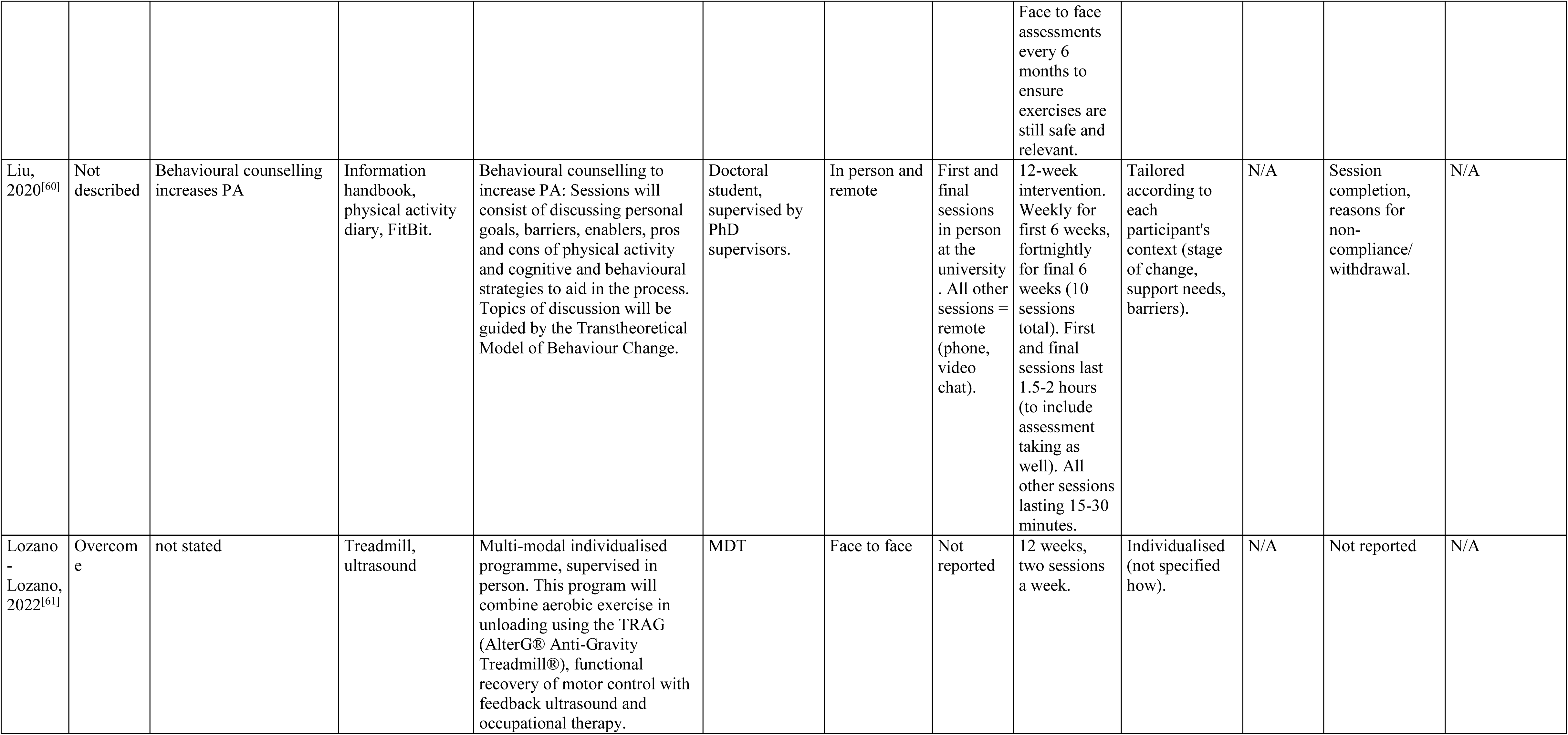
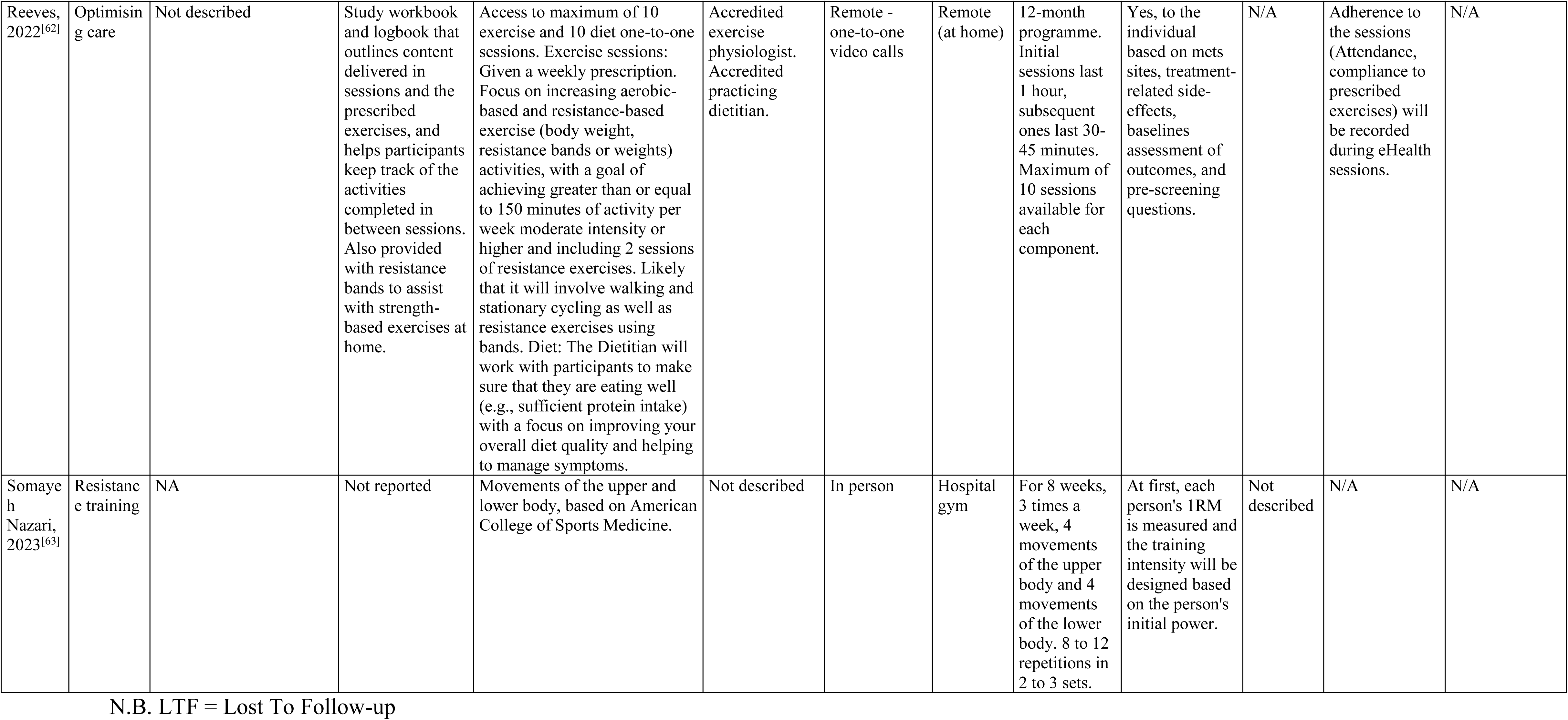

## Data Availability

All data produced in the present work are contained in the manuscript and/or available online at DOI 10.17605/OSF.IO/2SW76

https://osf.io/2sw76/

---

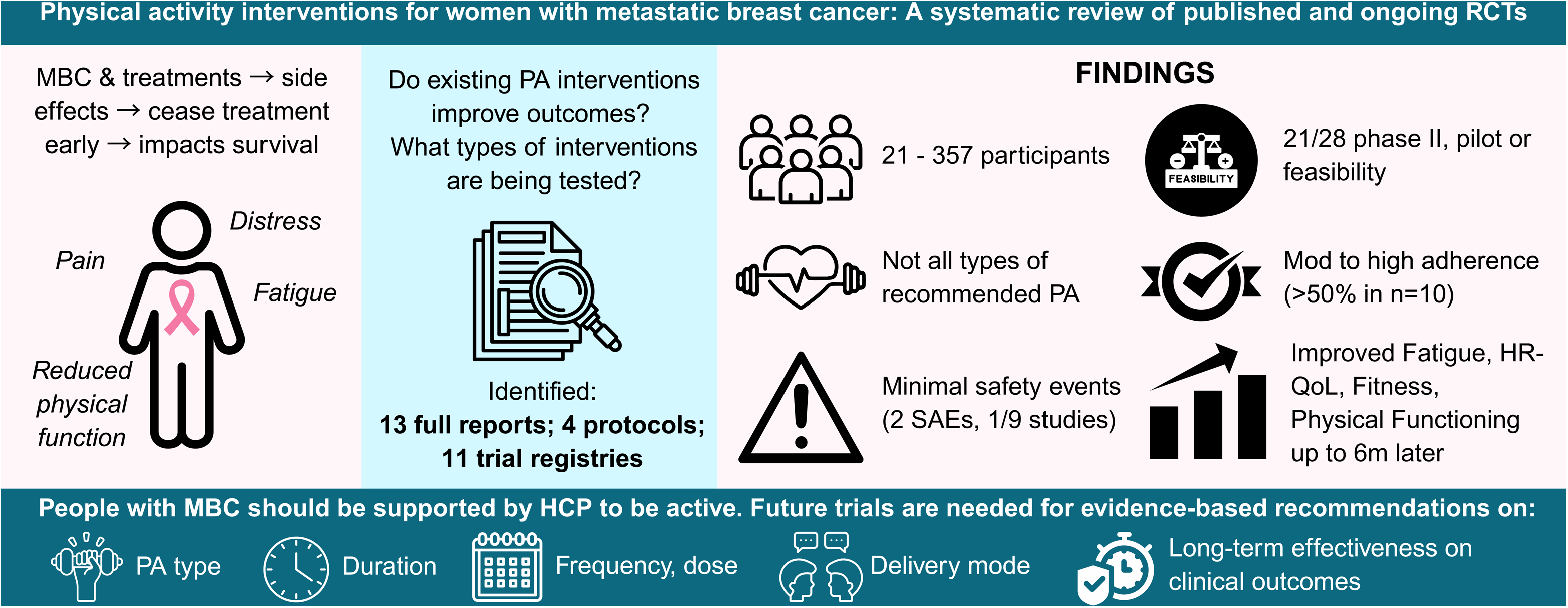

## References

1. Caswell-Jin JL, Plevritis SK, Tian L, et al. Change in Survival in Metastatic Breast Cancer with Treatment Advances: Meta-Analysis and Systematic Review. JNCI Cancer Spectr. 2018;2(4):pky062. doi:10.1093/jncics/pky062

2. Gogate A, Wheeler SB, Reeder-Hayes KE, et al. Projecting the Prevalence and Costs of Metastatic Breast Cancer From 2015 through 2030. JNCI Cancer Spectrum. 2021;5(4):pkab063. doi:10.1093/jncics/pkab063

3. Cancer survival in England - adults diagnosed - Office for National Statistics. Accessed November 22, 2022. https://www.ons.gov.uk/peoplepopulationandcommunity/healthandsocialcare/conditionsanddiseases/datasets/cancersurvivalratescancersurvivalinenglandadultsdiagnosed

4. Cardoso F, Spence D, Mertz S, et al. Global analysis of advanced/metastatic breast cancer: Decade report (2005-2015). Breast. 2018;39:131–138. doi:10.1016/j.breast.2018.03.002

5. Mosher CE, DuHamel KN. An examination of distress, sleep, and fatigue in metastatic breast cancer patients. Psycho-Oncology. 2012;21(1):100–107. doi:10.1002/pon.1873

6. Naik H, Leung B, Laskin J, et al. Emotional distress and psychosocial needs in patients with breast cancer in British Columbia: younger versus older adults. Breast Cancer Res Treat. 2020;179(2):471–477. doi:10.1007/s10549-019-05468-6

7. Fayanju OM, Yenokyan K, Ren Y, et al. The effect of treatment on patient-reported distress after breast cancer diagnosis. Cancer. 2019;125(17):3040–3049. doi:10.1002/cncr.32174

8. Mierzynska J, Taye M, Pe M, et al. Reference values for the EORTC QLQ-C30 in early and metastatic breast cancer. European Journal of Cancer. 2020;125:69–82. doi:10.1016/j.ejca.2019.10.031

9. Reed E, Simmonds P, Haviland J, Corner J. Quality of Life and Experience of Care in Women With Metastatic Breast Cancer: A Cross-Sectional Survey. Journal of Pain and Symptom Management. 2012;43(4):747–758. doi:10.1016/j.jpainsymman.2011.05.005

10. Yee J, Davis GM, Beith JM, et al. Physical activity and fitness in women with metastatic breast cancer. J Cancer Surviv. 2014;8(4):647–656. doi:10.1007/s11764-014-0378-y

11. Walker MS, Masaquel AS, Kerr J, et al. Early treatment discontinuation and switching in first-line metastatic breast cancer: the role of patient-reported symptom burden. Breast Cancer Res Treat. 2014;144(3):673–681. doi:10.1007/s10549-014-2892-z

12. McNeely ML, Campbell KL, Rowe BH, Klassen TP, Mackey JR, Courneya KS. Effects of exercise on breast cancer patients and survivors: a systematic review and meta-analysis. CMAJ. 2006;175(1):34–41. doi:10.1503/cmaj.051073

13. Ligibel JA, Bohlke K, May AM, et al. Exercise, Diet, and Weight Management During Cancer Treatment: ASCO Guideline. JCO. 2022;40(22):2491–2507. doi:10.1200/JCO.22.00687

14. Campbell KL, Winters-Stone K, Wiskemann J, et al. Exercise Guidelines for Cancer Survivors: Consensus statement from International Multidisciplinary Roundtable. Med Sci Sports Exerc. 2019;51(11):2375–2390. doi:10.1249/MSS.0000000000002116

15. Heywood R, McCarthy AL, Skinner TL. Safety and feasibility of exercise interventions in patients with advanced cancer: a systematic review. Support Care Cancer. 2017;25(10):3031–3050. doi:10.1007/s00520-017-3827-0

16. De Lazzari N, Niels T, Tewes M, Goette M. A systematic review of the safety, feasibility and benefits of exercise for patients with advanced cancer. Cancers. 2021 Sep 6;13(17):4478. doi:10.3390/cancers13174478

17. Campbell KL, Cormie P, Weller S, et al. Exercise Recommendation for People With Bone Metastases: Expert Consensus for Health Care Providers and Exercise Professionals. JCO Oncol Pract. 2022;18(5):e697–e709. doi:10.1200/OP.21.00454

18. Hart NH, Poprawski DM, Ashbury F, et al. Exercise for people with bone metastases: MASCC endorsed clinical recommendations developed by the International Bone Metastases Exercise Working Group. Support Care Cancer. 2022;30(9):7061–7065. doi:10.1007/s00520-022-07212-1

19. Weller S, Hart NH, Bolam KA, et al. Exercise for individuals with bone metastases: A systematic review. Crit Rev Oncol Hematol. 2021;166:103433. doi:10.1016/j.critrevonc.2021.103433

20. Lee CY, Laffoon K, Mama SK, et al. Outcomes for breast cancer survivors with metastatic disease in a physical activity program for medically underserved cancer survivors. J Cancer Surviv. Published online May 27, 2024. doi:10.1007/s11764-024-01600-8

21. Caterino JM, Adler D, Durham DD, et al. Analysis of Diagnoses, Symptoms, Medications, and Admissions Among Patients With Cancer Presenting to Emergency Departments. JAMA Netw Open. 2019;2(3):e190979. doi:10.1001/jamanetworkopen.2019.0979

22. Schmidt ME, Scherer S, Wiskemann J, Steindorf K. Return to work after breast cancer: The role of treatment-related side effects and potential impact on quality of life. Eur J Cancer Care (Engl). 2019;28(4):e13051. doi:10.1111/ecc.13051

23. de Boer AG, Taskila T, Tamminga SJ, Frings-Dresen MH, Feuerstein M, Verbeek JH. Interventions to enhance return-to-work for cancer patients. Cochrane Database Syst Rev. 2011;(2):CD007569. doi:10.1002/14651858.CD007569.pub2

24. Svensson H, Hatschek T, Johansson H, Einbeigi Z, Brandberg Y. Health-related quality of life as prognostic factor for response, progression-free survival, and survival in women with metastatic breast cancer. Med Oncol. 2012;29(2):432–438. doi:10.1007/s12032-011-9844-9

25. Efficace F, Biganzoli L, Piccart M, et al. Baseline health-related quality-of-life data as prognostic factors in a phase III multicentre study of women with metastatic breast cancer. European Journal of Cancer. 2004;40(7):1021–1030. doi:10.1016/j.ejca.2004.01.014

26. Delrieu L, Jacquet E, Segura-Ferlay C, et al. Analysis of the StoRM cohort reveals physical activity to be associated with survival in metastatic breast cancer. Sci Rep. 2020;10(1):10757. doi:10.1038/s41598-020-67431-6

27. Toohey K, Chapman M, Rushby AM, Urban K, Ingham G, Singh B. The effects of physical exercise in the palliative care phase for people with advanced cancer: a systematic review with meta-analysis. J Cancer Surviv. 2023;17(2):399–415. doi:10.1007/s11764-021-01153-0

28. Rodríguez-Cañamero S, Cobo-Cuenca AI, Carmona-Torres JM, et al. Impact of physical exercise in advanced-stage cancer patients: Systematic review and meta-analysis. Cancer Med. 2022;11(19):3714–3727. doi:10.1002/cam4.4746

29. Dittus KL, Gramling RE, Ades PA. Exercise interventions for individuals with advanced cancer: A systematic review. Prev Med. 2017;104:124–132. doi:10.1016/j.ypmed.2017.07.015

30. Haider ZF, Beeken RJ, Ayaz-Shah A, Lloyd KE, Smith SG. ‘As Important as Medication’. A Qualitative Investigation of the Beliefs, Barriers and Facilitators of Physical Activity for Women With Metastatic Breast Cancer. Psycho-Oncology. 2025 Jun;34(6):e70193. doi:10.1002/pon.70193

31. Aromataris E, Munn Z. JBI Manual for Evidence Synthesis. Jbi; 2020. 10.46658/JBIMES-20-01

32. Page MJ, McKenzie JE, Bossuyt PM, et al. The PRISMA 2020 statement: an updated guideline for reporting systematic reviews. BMJ. 2021;372:n71. doi:10.1136/bmj.n71

33. Tufanaru C, Munn Z, Aromataris E, Campbell J, Hopp L. Chapter 3: Systematic reviews of effectiveness. In: Aromataris E, Munn Z, eds. Joanna Briggs Institute Reviewer’s Manual. Vol 3. The Joanna Briggs Institute Adelaide, Australia; 2017. Accessed June 3, 2024. https://reviewersmanual.joannabriggs.org/

34. Hoffmann TC, Glasziou PP, Boutron I, Milne R, Perera R, Moher D, Altman DG, Barbour V, Macdonald H, Johnston M, Lamb SE. Better reporting of interventions: template for intervention description and replication (TIDieR) checklist and guide. Bmj. 2014 Mar 7;348. doi: 10.1136/bmj.g1687

35. Physical activity. Accessed June 3, 2024. https://www.who.int/news-room/fact-sheets/detail/physical-activity

36. The 4 most important types of exercise. Harvard Health. January 13, 2017. Accessed June 3, 2024. https://www.health.harvard.edu/exercise-and-fitness/the-4-most-important-types-of-exercise

37. Baldwin JN, Hassett L, Sherrington C. Framework to Classify Physical Activity Intervention Studies for Older Adults. Translational Journal of the American College of Sports Medicine. 2023;8(3):e000230. doi:10.1249/TJX.0000000000000230

38. Goldschmidt S, Schmidt ME, Steindorf K. Long-term effects of exercise interventions on physical activity in breast cancer patients: a systematic review and meta-analysis of randomized controlled trials. Support Care Cancer. 2023;31(2):130. doi:10.1007/s00520-022-07485-6

39. Carson JW, Carson KM, Olsen M, et al. Yoga Practice Predicts Improvements in Day-to-Day Pain in Women with Metastatic Breast Cancer. J Pain Symptom Manage. 2021;61(6):1227–1233. doi:10.1016/j.jpainsymman.2020.10.009

40. Headley JA, Ownby KK, John LD. The effect of seated exercise on fatigue and quality of life in women with advanced breast cancer. Oncol Nurs Forum. 2004;31(5):977–983. doi:10.1188/04.ONF.977-983

41. Ligibel JA, Giobbie-Hurder A, Shockro L, et al. Randomized trial of a physical activity intervention in women with metastatic breast cancer. Cancer. 2016;122(8):1169–1177. doi:10.1002/cncr.29899

42. Oh B, Butow PN, Boyle F, et al. Effects of qigong on quality of life, fatigue, stress, neuropathy, and sexual function in women with metastatic breast cancer: A feasibility study. International Journal of Physical anret nI Medicine & Rehabilitation. 2014;2(4). doi:10.4172/2329-9096.1000217

43. Porter LS, Carson JW, Olsen M, et al. Feasibility of a mindful yoga program for women with metastatic breast cancer: results of a randomized pilot study. Support Care Cancer. 2019;27(11):4307–4316. doi:10.1007/s00520-019-04710-7

44. Schmitz KH, Kanski B, Gordon B, et al. Technology-based supportive care for metastatic breast cancer patients. Support Care Cancer. 2023;31(7):401. doi:10.1007/s00520-023-07884-3

45. Schmitz KH, Schleicher E, Doerksen S, et al. Testing the acceptability and feasibility of a tablet-based supportive cancer platform for patients with metastatic breast cancer. J Cancer Surviv. 2021;15(3):410–413. doi:10.1007/s11764-021-01021-x

46. Scott JM, Iyengar NM, Nilsen TS, et al. Feasibility, safety, and efficacy of aerobic training in pretreated patients with metastatic breast cancer: A randomized controlled trial. Cancer. 2018;124(12):2552–2560. doi:10.1002/cncr.31368

47. Sheean P, Matthews L, Visotcky A, et al. Every Day Counts: a randomized pilot lifestyle intervention for women with metastatic breast cancer. Breast Cancer Res Treat. 2021;187(3):729–741. doi:10.1007/s10549-021-06163-1

48. Vadiraja H, Rao RM, Nagarathna R, et al. Effects of Yoga in Managing Fatigue in Breast Cancer Patients: A Randomized Controlled Trial. Indian J Palliat Care. 2017;23(3):247–252. doi:10.4103/IJPC.IJPC_95_17

49. Yee J, Davis GM, Hackett D, et al. Physical Activity for Symptom Management in Women With Metastatic Breast Cancer: A Randomized Feasibility Trial on Physical Activity and Breast Metastases. J Pain Symptom Manage. 2019;58(6):929–939. doi:10.1016/j.jpainsymman.2019.07.022

50. Hiensch AE, Depenbusch J, Schmidt ME, et al. Supervised, structured and individualized exercise in metastatic breast cancer: a randomized controlled trial. Nat Med. 2024;30(10):2957–2966. doi:10.1038/s41591-024-03143-y

51. Phillips SM, Starikovsky J, Solk P, et al. Feasibility and preliminary effects of the Fit2ThriveMB pilot physical activity promotion intervention on physical activity and patient reported outcomes in individuals with metastatic breast cancer. Breast Cancer Res Treat. 2024;208(2):391–403. doi:10.1007/s10549-024-07432-5

52. Delrieu L, Anota A, Trédan O, et al. Design and methods of a national, multicenter, randomized and controlled trial to assess the efficacy of a physical activity program to improve health-related quality of life and reduce fatigue in women with metastatic breast cancer: ABLE02 trial. BMC Cancer. 2020;20(1):622. doi:10.1186/s12885-020-07093-9

53. Hart NH, Galvão DA, Saunders C, et al. Mechanical suppression of osteolytic bone metastases in advanced breast cancer patients: a randomised controlled study protocol evaluating safety, feasibility and preliminary efficacy of exercise as a targeted medicine. Trials. 2018;19(1):695. doi:10.1186/s13063-018-3091-8

54. Kirkham AA, King K, Joy AA, et al. Rationale and design of the Diet Restriction and Exercise-induced Adaptations in Metastatic breast cancer (DREAM) study: a 2-arm, parallel-group, phase II, randomized control trial of a short-term, calorie-restricted, and ketogenic diet plus exercise during intravenous chemotherapy versus usual care. BMC Cancer. 2021;21(1):1093. doi:10.1186/s12885-021-08808-2

55. Meier-Girard D, Ribi K, Gerstenberg G, Ruhstaller T, Wolf U. Eurythmy therapy versus slow movement fitness in the treatment of fatigue in metastatic breast cancer patients: study protocol for a randomized controlled trial. Trials. 2020;21(1):612. doi:10.1186/s13063-020-04542-5

56. NCT05468034. Exercise in Metastatic Breast Cancer: eMBody. https://clinicaltrials.gov/show/NCT05468034. Published online August 31, 2022. doi:10.1002/central/CN-02424684

57. NCT01653366. Effect of a Goal Setting Intervention on Exercise in Women With Metastatic Breast Cancer: an Exploration of The Lipogenic Pathway. https://clinicaltrials.gov/show/NCT01653366. Published online May 31, 2018. doi:10.1002/central/CN-01585158

58. ACTRN12614000350628. Randomised study to evaluate the impact of Aerobic and Resistance Exercise on fatigue in patients with advanced breast cancer. https://trialsearch.who.int/Trial2.aspx?TrialID=ACTRN12614000350628. Published online March 31, 2019. doi:10.1002/central/CN-01874124

59. ISRCTN - ISRCTN12841416: Breast activity and healthy eating after diagnosis 3. doi:10.1186/ISRCTN12841416

60. ACTRN12620000743965. Behaviour Change for Physical Activity after Metastatic Breast Cancer. https://trialsearch.who.int/Trial2.aspx?TrialID=ACTRN12620000743965. Published online October 31, 2020. doi:10.1002/central/CN-02165426

61. NCT05244382. Overcome, a Program of Therapeutic Exercise and Functional Recovery to Improve the Functional Capacity of Women With Breast Cancer and Bone Metastases. https://clinicaltrials.gov/show/NCT05244382. Published online March 31, 2022. doi:10.1002/central/CN-02381619

62. ACTRN12622001494729. Optimising Care: supporting women with metastatic breast cancer to optimise their quality of life via exercise and diet. https://trialsearch.who.int/Trial2.aspx?TrialID=ACTRN12622001494729. Published online December 31, 2022. doi:10.1002/central/CN-02503064

63. IRCT20200208046418N2. The effect of exercise on metastatic breast cancer patients. https://trialsearch.who.int/Trial2.aspx?TrialID=IRCT20200208046418N2. Published online June 30, 2023. doi:10.1002/central/CN-02566324

64. NCT05277935. Wearable Enhanced Fitness Tracking for Metastatic Breast Cancer Patients Using Endocrine Treatment and Palbociclib. https://clinicaltrials.gov/show/NCT05277935. Published online March 31, 2022. doi:10.1002/central/CN-02382414

65. Study Details | Prolonged Overnight Fasting and/or Exercise on Fatigue and Other Patient Reported Outcomes in Women With Hormone Receptor Positive Advanced Breast Cancer | ClinicalTrials.gov. Accessed October 1, 2024. https://clinicaltrials.gov/study/NCT06123988

66. Study Details | The FEMA Study: Feasibility of Exercise in Patients With Metastatic Breast Cancer and Adiposity | ClinicalTrials.gov. Accessed October 1, 2024. https://clinicaltrials.gov/study/NCT06343987

67. Sweegers MG, Depenbusch J, Aaronson NK, Hiensch AE, Wengström Y, Backman M, Gunasekara N, Clauss D, Belloso J, Lachowicz M, May AM. Metastatic breast cancer patients’ preferences for exercise programs: a latent class analysis using data from a survey in five European countries. Supportive Care in Cancer. 2025 Jan;33(1):39. doi:10.1007/s00520-024-09068-z

68. Collado-Mateo D, Lavín-Pérez AM, Peñacoba C, Del Coso J, Leyton-Román M, Luque-Casado A, Gasque P, Fernandez-del-Olmo MA, Amado-Alonso D. Key factors associated with adherence to physical exercise in patients with chronic diseases and older adults: an umbrella review. International journal of environmental research and public health. 2021 Feb;18(4):2023. doi:10.3390/ijerph18042023

69. Klompstra L, Deka P, Almenar L, Pathak D, Munoz-Gomez E, López-Vilella R, Marques-Sule E. Physical activity enjoyment, exercise motivation, and physical activity in patients with heart failure: A mediation analysis. Clinical Rehabilitation. 2022 Oct;36(10):1324–31. doi10.1177/02692155221103

70. Skivington K, Matthews L, Simpson SA, et al. A new framework for developing and evaluating complex interventions: update of Medical Research Council guidance. BMJ. 2021;374:n2061. doi:10.1136/bmj.n2061

71. Carver CS, Scheier MF. Control theory: A useful conceptual framework for personality–social, clinical, and health psychology. Psychological bulletin. 1982 Jul;92(1):111. doi:10.1037/0033-2909.92.1.111

72. Michie S, Abraham C, Whittington C, McAteer J, Gupta S. Effective techniques in healthy eating and physical activity interventions: a meta-regression. Health psychology. 2009 Nov;28(6):690. doi: 10.1037/a0016136

73. Michie S, Richardson M, Johnston M, et al. The behavior change technique taxonomy (v1) of 93 hierarchically clustered techniques: building an international consensus for the reporting of behavior change interventions. Ann Behav Med. 2013;46(1):81–95. doi:10.1007/s12160-013-9486-6

74. Marques MM, Wright AJ, Corker E, et al. The Behaviour Change Technique Ontology: Transforming the Behaviour Change Technique Taxonomy v1. Wellcome Open Res. 2023;8:308. doi:10.12688/wellcomeopenres.19363.1

75. Collins LM, Strayhorn JC, Vanness DJ. One view of the next decade of research on behavioral and biobehavioral approaches to cancer prevention and control: intervention optimization. Translational Behavioral Medicine. 2021;11(11):1998–2008. doi:10.1093/tbm/ibab087

76. Michie S, Johnston M, Francis J, Hardeman W, Eccles M. From Theory to Intervention: Mapping Theoretically Derived Behavioural Determinants to Behaviour Change Techniques. Applied Psychology. 2008;57(4):660–680. doi:10.1111/j.1464-0597.2008.00341.x

